# Strategies to minimize SARS-CoV-2 transmission in classroom settings: Combined impacts of ventilation and mask effective filtration efficiency

**DOI:** 10.1101/2020.12.31.20249101

**Authors:** David A. Rothamer, Scott Sanders, Douglas Reindl, Timothy H. Bertram

## Abstract

The impact of the COVID-19 pandemic continues to be significant and global. As the global community learns more about the novel coronavirus SARS-CoV-2, there is strong evidence that a significant modality of transmission is via the long-range airborne route, referred to here as aerosol transmission. In this paper, we evaluate the efficacy of ventilation, mask effective filtration efficiency, and the combined effect of the two on the reduction of aerosol infection probability for COVID-19 in a classroom setting. The Wells-Riley equation is used to predict the conditional probability of infection for three distinct airborne exposure scenarios: (1) an infectious instructor exposing susceptible students; (2) an infectious student exposing other susceptible students; and (3) an infectious student exposing a susceptible instructor. Field measurements were performed in a classroom using a polydisperse neutralized salt (NaCl) aerosol, generated in a size range consistent with human-generated SARS-CoV-2 containing bioaerosols, as a safe surrogate. Measurements included time-resolved and size-resolved NaCl aerosol concentration distributions and size-resolved effective filtration efficiency of different masks with and without mask fitters. The measurements were used to validate assumptions and inputs for the Wells-Riley model. Aerosol dynamics and distribution measurements confirmed that the majority of the classroom space is uniform in aerosol concentration within a factor of 2 or better for distances *>* 2 m from the aerosol source. Mask effective filtration efficiency measurements show that most masks fit poorly with estimated leakage rates typically *>* 50%, resulting in significantly reduced effective filtration efficiency. However, effective filtration efficiencies approaching the mask material filtration efficiency were achievable using simple mask fitters. Wells-Riley model results for the different scenarios suggest that ventilation of the classroom alone is not able to achieve infection probabilities less than 0.01 (1%) for air exchanges rates up to 10 *h^−^*^1^ and an event duration of one hour. The use of moderate to high effective filtration efficiency masks by all individuals present, on the other hand, was able to significantly reduce infection probability and could achieve reductions in infection probability by 5x, 10x, or even *>*100x dependent on the mask used and use of a mask fitter. This enables conditional infection probabilities *<* 0.001 (0.1%) or even *<* 0.0001 (0.01%) to be reached with the use of masks and mask fitters alone. Finally, the results demonstrate that the reductions provided by ventilation and masks are synergistic and multiplicative. The results reinforce the use of properly donned masks to achieve reduced aerosol transmission of SARS-CoV-2 and other infectious diseases transmitted via respiratory aerosol indoors and provide new motivation to further improve the effective filtration efficiency of common face coverings through improved design, and/or the use of mask fitters.

## 1 Introduction

Foundational to infection control and prevention is a clear understanding of the modality of transmission of the contagion. For SARS-CoV-2, early during the COVID-19 pandemic, emphasis was placed on direct-contact and indirect-contact modes of transmission with recommendations focused on hand hygiene [1] and physical distancing [2]; however, recognition of the long-range airborne route as a modality of transmission [3–6] means that appropriate intervention measures for infection control and prevention via airborne transmission are urgently needed. A common approach for assessing long-range airborne transmission, referred to here as aerosol transmission, utilizes the Wells-Riley equation [7, 8] to relate aerosol concentrations to infection probability assuming an exponential dose model [9]. This approach has been applied widely for SARS-CoV-2, and other diseases [8, 9] for both prospective [10] and retrospective analyses [10, 11], with a particular focus on super-spreader events [11, 12].

The application of the Wells-Riley equation to a simple well-mixed room control-volume model (often termed a box model) is useful for risk assessment and planning of interventions to reduce the risk of disease transmission. Use of this approach requires information on the setting of the event (room and heating ventilation and air conditioning (HVAC) system information), event duration, the potential emission rate of infectious aerosol and breathing rates of susceptible individuals, and information on interventions being considered. Of these parameters, emission rate of infectious aerosol tends to be the most uncertain, although recent work has better defined the range of values to consider as a function of activity level and expiratory activity for SARS-CoV-2 [10, 13]. The next most uncertain parameters are the room ventilation rate and removal rate of virus in the room via either HVAC system or other common interventions such as masks. In this work, we assume that measures have already been put in place to reduce occupant density in the room to control short-range droplet transmission and controls have been put in place to control fomite transmission. The focus of this paper is on the direct measurement of input parameters for the well-mixed Wells-Riley model related to interventions to reduce infection risk via aerosol transmission in a classroom setting. This includes assessment of HVAC flow rates and aerosol decay rate in a university classroom and measurements of mask filtration efficiency for four different types of masks. The effectiveness of “mask fitters” designed to seal masks to the user’s face is also evaluated. One of the aims of this paper is to provide recommendations that, when implemented, can decrease the likelihood of COVID-19 transmission in traditionally high occupancy spaces such as classrooms.

## 2 Background

### 2.1 Aerosol particles and airborne modality of transmission of COVID-19

As Nardell and Nathavitharana [5] note, the term airborne and the related lexicon as they apply to infectious bioaerosols have not been uniformly defined in the context of person-to-person spread of viral respiratory pathogens. Here, we define a respiratory particle as an aqueous droplet comprised of saliva, mucous, infectious agent(s), and other biomatter. Respiratory particles can be generated directly and dispersed to the air by a person breathing, coughing, sneezing, talking, singing, or vomiting and indirectly by aerosolizing feces during sewage removal and treatment [14]. When an infected individual is actively shedding viruses, these dispersed respiratory particles serve as a transport vehicle with each respiratory particle potentially carrying a viral payload into the air.

The size of respiratory particles can range from submicron to hundreds of microns in aerody-namic diameter. Johnson et al. [15] identified three modes in the distribution of particles resulting from human expiratory activities: the bronchiolar fluid film burst (B) mode originating in the lungs due to breathing (mode median diameter 1.6 *µ*m); the laryngeal (L) mode associated with the larynx, active during vocalization (mode median diameter 1.7-2.5 *µ*m); and the oral (O) cavity mode (mode median diameter 123-145 *µ*m) active during speech and coughing. Large droplets generated in the mouth via the oral mode (greater than *∼*100 *µ*m in diameter) tend to quickly fall to the ground by gravity and remain within about a 2.0 m distance from the generation source when emitted due breathing or coughing [16]. It should be noted that during sneezing, large droplets can be expelled to much greater distances due to the high velocity jet generated [17]. If these large respiratory droplets are inhaled, they tend to be deposited in the upper respiratory tract [18] Aerosol particles generated by breathing, vocalization, and coughing due to the B and L modes and fall in the 200 nm – 10 *µ*m size range. Particles smaller than 10 *µ*m are easily inhaled and able to penetrate deeper into the respiratory tract [19], depositing deep in the lungs.

Our focus in this work is on inhalable virus laden aerosol particles (bioaerosols) in the 100 nm – 5 *µ*m size range for several reasons. First, aerosol particles in this size range have been demonstrated to carry viable SARS-CoV-2 [20]. Second, they are consistent with the particle sizes humans generate by common activities in a classroom that include breathing, speaking, singing, coughing, and sneezing [14, 15, 21]. Third, aerosol particles in this size range readily breach the current 2-m physical-distancing guidelines because of their ability to remain airborne for extended periods of time and be readily carried by air currents in the indoor environment. Respiratory droplets less than 10 *µ*m in size rapidly (*<* 1 s) reach an equilibrium diameter about one-half their original diameter due to evaporation and equilibration at the ambient conditions [21–24]. Therefore, the size range considered corresponds to droplets at the point of emission ranging in size from 200 nm to 10 *µ*m and to an equilibrium particle size range from 100 nm to 5 *µ*m consistent with the size range described by Fennelly [17].

Beyond recommendations aiming to limit the spread of COVID-19 in indoor spaces that include hand hygiene, cleaning/sanitizing solid surfaces, physical distancing, and similar measures, the United States Centers for Disease Control and Prevention (CDC) also recommend (as of 12/13/2020) [25, 26]:

- Occupants should use non-medical disposable or cloth masks. The CDC excludes medical masks (procedure and surgical masks) and N95 respirators in order to conserve those for healthcare workers.
- Ventilating spaces to the greatest extent possible, including increasing circulation rate of outdoor air.

The current work seeks to better understand the efficacy of these interventions in terms of the decrease in probability of infection due to aerosol transmission which they provide in a classroom setting.

### 2.2 Estimating probability of COVID-19 infection by aerosols containing SARS-CoV-2

In 1974, a measles outbreak occurred in an elementary school in upstate New York. The index case was a second grader who produced 28 secondary cases in 14 different classrooms within the school and two subsequent generations yielded 60 infections. This case was analyzed by Riley et al. [8] and, as part of their analysis, they proposed a mathematical model to predict the probability of infection via airborne route based on individual characteristics such as breathing rate and building HVAC characteristics such as room ventilation rate. The Wells-Riley equation [8] predicts the conditional probability of infection, *P*, based on a susceptible individual inhaling an infectious quanta dose, *D_q_*, during the duration of an event given by

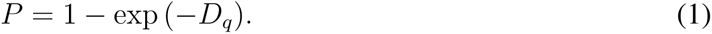

A single infectious quantum dose is related to the number of bioaerosol particles (and number of viable virus copies) inhaled that results in a conditional probability of infection of *P* = 1 *−* exp (*−*1) = 63.2%.

The efficacy of the equation at predicting infection probability is based on how well the underlying assumptions hold. These assumption include [9]:

1. Virus incubation period *»* timescale for the event,
2. Single large dose is equivalent to multiple small doses over a period of time, and
3. Probabilistic approach, best suited to large populations.

The first two assumptions may hold well for many scenarios where the equation is applied for SARS-CoV-2 in high-risk settings where exposure has a limited duration. The third assumption is related to the limits of deterministic application of the model to situations with small numbers of individuals.

Estimation of infection probability due to airborne transmission requires calculation of the evolution of bioaerosol concentration in the space of interest in order to estimate the number of infectious quanta a susceptible individual inhales. The Wells-Riley equation is typically utilized by applying a well-mixed room approximation using a simple control-volume model, as was done by Riley et al. [8]. In the case of an infectious individual(s) present in the room, the concentration we are interested in is the concentration of infectious aerosol, for measurements we present in the paper regarding the efficacy of different interventions, we seed NaCl aerosol into the room to perform evaluations of aerosol dynamics and distribution and mask filtration efficiency. In that case, we are interested in describing the evolution of the concentration of NaCl particles in the room.

#### 2.2.1 Room aerosol concentration time evolution

The aerosol concentration in a room where particles are being supplied inside the room (either by an infectious individual or by another aerosol source), assuming the that room is perfectly mixed, is described by the first order ordinary differential equation (similar to the material balance approach of Nazaroff [27])

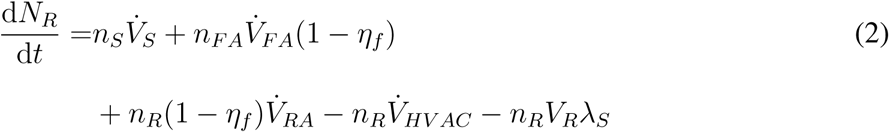

where *N_R_* is the number of aerosol particles in the room, *n_S_* is the number density (number concentration) of aerosol particles in the aerosol being supplied inside the room, 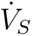 is volume flowrate of aerosol being supplied inside the room, *n_FA_* is the number density of particles in the fresh air, 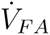 is the volumetric flowrate of fresh outdoor air, *η_f_* is the filtration efficiency of the HVAC system, *n_R_* is the number density of particles in the room, 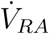 is the volumetric flowrate of recycled air, 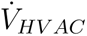 is the total HVAC flowrate (total return flowrate), *V_R_* is the room volume, and *λ_S_* is the total first order loss rate due to settling in the room (given by *λ_S_* = *V_TS_/H* where *V_TS_* is the terminal settling velocity and *H* is the height from which particles settle). Here we have neglected dilution effects due to natural ventilation and infiltration.

Dividing by the volume of the room allows Equation 2 to be converted to an ODE for the number density of particles in the room, i.e,

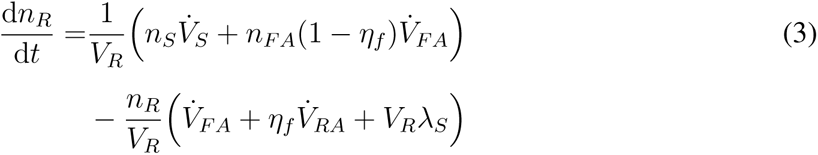

where we have used 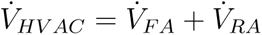. Defining

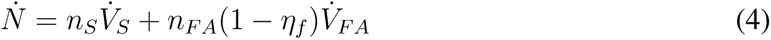

as the total external supply rate of particles and

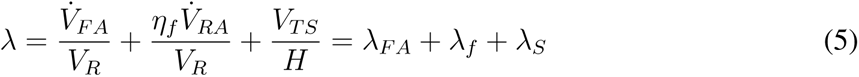

as the total loss rate of particles, the ODE can be rewritten as

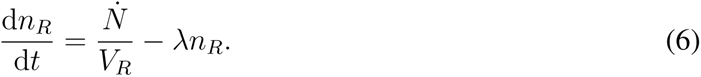

Solving the ODE for *n_R_* assuming an initial number density of particles in the room *n_R_*_0_ gives

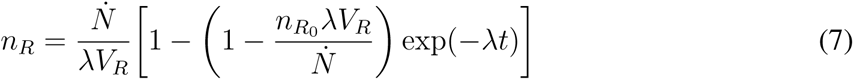

which provides the time-dependent concentration of particles in the room. It should be noted that this result could be applied in a size-dependent fashion by solving the equation for different size bins using size-dependent filtration efficiencies and terminal settling velocities. Otherwise, if used for total concentration, weighted average values for *η_f_* and *V_TS_* should be used. At long times (*λt »* 1), a steady-state concentration (*n_R,ss_*) is reached

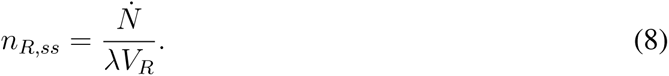

#### 2.2.2 Infectious quanta number density and dose

In the case of an infectious individual for the Wells-Riley equation, the result in Equation 7 can be rewritten in terms of the total net emission rate of infectious quanta 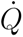 to determine the number density of infectious quanta in the room (*n_q_*) as a function of time, i.e,

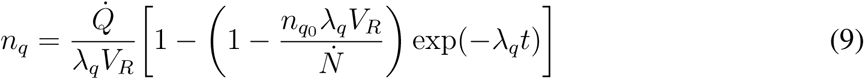

where *λ_q_* is now the total first order loss rate for quanta in the room. The net total emission rate of quanta 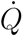 is related to the individual quanta emission rate 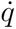 by

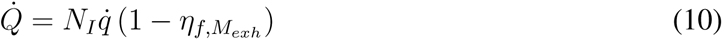

where *N_I_* is the number of infectious individuals in the room (usually assumed to be 1) and *η_f,Mexh_* is the effective filtration efficiency of masks worn by individuals in the room during exhalation. The net total quanta emission rate defined in this way enables the use of masks to be explicitly included in the analysis.

The infectious quantum dose, *D_q_*, is determined by multiplying the average number density of quanta in the room (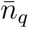) during an event and the breathing rate of susceptibles (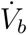) in the room by the duration of the event (*t_D_*), i.e,

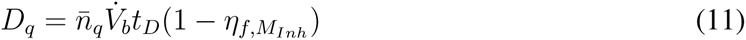

where number of quanta inhaled (dose) is written to take into account the effective filtration efficiency during inhalation (*η_f,MInh_*) of masks worn by susceptible individuals in the room. In the current study, the infectious quanta is assumed to be SARS-CoV-2.

The average concentration of quanta in the room (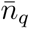) is determined from integration of Equation 9 over the time period of interest. For the simplified case with no infectious quanta initially present in the room at the start of the event duration of interest, integration of Equation 9 gives

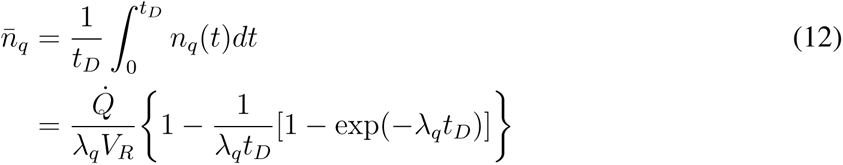

where *λ_q_* is a first-order loss rate coefficient comprised of the various mechanisms that are responsible for “removing” infectious quanta from the room volume. This includes the mechanisms discussed earlier for particle removal, but also includes an additional loss term for virus inactivation (*k*), i.e.,

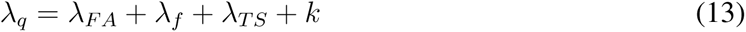

where *λ_FA_* represents the dilution effect of fresh air, *λ_f_* accounts for the removal of quanta due to filtration (central and in-room systems would be additive, as applicable), *λ_TS_* is the loss due to quanta settling in the room or HVAC system deposition, and *k* represents the rate of inactivation of the viral quanta. Additional loss mechanisms that may be applicable to a given situation can be added to Equation 13.

To more easily see the dependence of infection probability on the parameters of interest, it is useful to look at an approximate expression for a limiting case. For example, if *D_q_* in Equation 1 is small (i.e., *D_q_ ≤* 0.2) then exp(*−D_q_*) *≈* 1 *− D_q_* and the probability of infection for small *D_q_* becomes

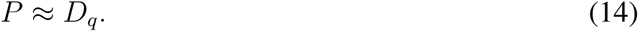

This holds within about 10% for conditional infection probabilities *≤* 20% which is consistent with the range we are interested in. Assuming a steady-state value for the quanta in the room and substituting the for *D_q_* and 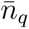 using Equations 11, 12, and 10 we can get an idea of the dependence of infection probability on the parameters of interest

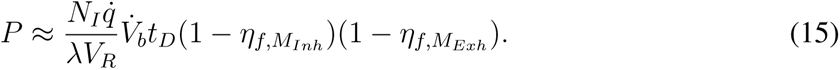

From Equation 15, we see that in the limit of small *D_q_* (i.e., relatively small infection probability), the conditional infection probability is proportional the quanta emission rate, the breathing rate of susceptibles, the duration of the exposure, and the product of the mask penetrations for inhalation and exhalation (penetration is equal to one minus the filtration efficiency [19]), and is inversely proportional to room volume and the first order loss rate (e.g., ventilation flow rate). It is important to note that if we assume the susceptible and infectious individuals are wearing the same mask, then we have a squared dependence on mask penetration (i.e., one minus mask filtration efficiency), this effect is stronger than the first order loss coefficient that can be increased by increasing room ventilation.

### 2.3 Room aerosol dynamics and distribution

The behavior of aerosols in indoor spaces has seen significant study due to the impact of particulates on human health (for examples see the reviews by Wallace [28], Narazoff [27], and Tham [29]). Emphasis has also been placed on the impacts of aerosol dynamics and distribution indoors and the potential for bioaerosols to result in aerosol transmission of diseases [30, 31]. Recently, a number of studies have focused on the dispersal of aerosols and dynamics of aerosol particle lifetimes in indoor spaces related to the spread of SARS-CoV-2 [24, 32–35].

It is well known from these, and other studies, that ventilation and air flow within a room impact aerosol concentration and transport [12, 27, 31, 32, 36]. To first order, the average concentration of aerosol in the room will be inversely proportional to the air-exchange rate in the room, as indicated in Equation 8. Room air-exchange rates vary significantly based on the type of building and specific location [27]. For buildings equipped with variable air volume systems, air exchange rates for a given space will vary with occupancy and room thermal load. The majority of residences and offices in the US have air-exchange rates *<* 3 air changes per hour (ACH) [27].

Although widely studied, there is still a lack of prior work that provide data on aerosol dynamics and distributions in the bioaerosol size range relevant for aerosol transmission of COVID-19. This information is needed to provide accurate inputs to the Wells-Riley model, and to assess the validity of the well-mixed approximation used in such models for cases with and without occupants wearing masks.

### 2.4 Mask effective filtration efficiency

It is now recognized that it is critical for the general public to wear masks in indoor environments and outdoors when physical distancing may not be achievable [37, 38]. Evidence from laboratory experiments by Ueki et al. [39] demonstrating the ability of masks to reduce virus transmission using the SARS-CoV-2 virus in a generated aerosol, and real world examples of masks preventing transmission of the virus when masks are consistently worn [40, 41], both strongly support the use of masks. However, the effectiveness of a mask will vary significantly based on the filtration efficiency of the materials it is composed of and the fit of the mask to the user’s face [42–44].

The filtration performance of materials for both homemade cloth masks and commercially produced masks (cloth masks, disposable non-medical masks, medical masks, and KN95 and N95 masks) has been studied both before the pandemic and during [43–53]. For more details, a recent thorough review of the literature on mask filtration measurements was published by Clase et al. [54].

The size range of the measurements, filtration conditions, and testing equipment and methods vary greatly between studies, making comparisons difficult. Additionally, there is a general lack of data at appropriate filtration conditions for normal breathing rates that is size resolved and covers the range of interest for human produced bioaerosols (from approximately 100 nm to 10 *µ*m), and that considers the impact of mask fit on the filtration results in a realistic setting. The study by van der Sande et al. [42] is notable in that it used human subjects and studied them during multiple activities in a realistic setting, but the size range of particles in the study only extended out to *∼* 1 *µ*m and the measurements were not size resolved. In the current work, we seek to help fill this gap by performing size-resolved effective filtration efficiency measurements with manikins simulating occupants in a classroom setting over a size range from 140 nm to 3.2 *µ*m in aerodynamic diameter.

### 2.5 Focus of this study

The predictive capability of the Wells-Riley equation is dependent on establishing accurate values of the various parameters that are component elements in the model. Field measurements were made in an actual classroom to assess, in part, the validity of assumptions embedded in the Wells-Riley model (i.e., the room is well-mixed), and the effective filtration efficiency of four different types of masks with and without the use of mask fitters. In the sections that follow, we first describe the classroom space and methods used for the experimental characterization of the classroom space and masks. Next, we characterize the behavior of the polydisperse NaCl aerosol dynamics and distribution in the space. The effective filtration efficiency of four different mask types are presented. With the inputs accurately determined, the paper concludes with an assessment of the protective measures of interest and their impact on conditional probability of infection using the Wells-Riley model.

## 3 Methods and materials

### 3.1 Classroom space studied

The classroom space that served as a testbed for the present study is shown (in its pre-COVID-19 layout) in Figure 1. The classroom measures 126.4 m^2^ with a ceiling height of 2.870 m yielding a gross room volume of 362.6 m^3^. The room is served by a central station variable air volume (VAV) air handling unit that supplies multiple spaces in the building. Each individual space within the building is equipped with one or more VAV boxes that modulate supply air to the room in response to load changes. Air delivery into the room is provided by four-way throw fixed area diffusers arranged in a rectangular layout. Because the central-station air handling unit’s maximum and minimum air flowrates (88,900 m^3^/h and 28,000 m^3^/h, respectively) are much larger than the air flowrate to an individual room, significant dilution of any aerosol from the room will occur prior to recirculation back to the space. A single overheard return is located near the rear corner of the classroom.

**Figure 1:**
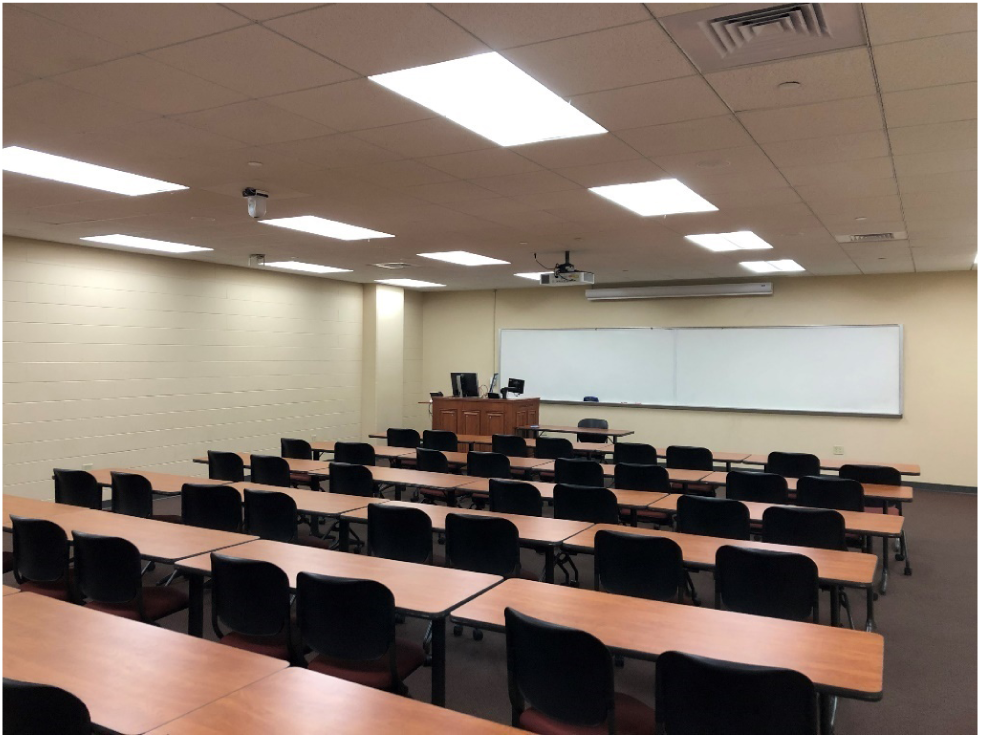
Photo showing the classroom space used as a testbed for the current study (in a pre-COVID-19 layout).

The minimum and maximum total air flowrates for the classroom space where testing was conducted ranged from 487.6 m^3^/h to 1,832 m^3^/h with corresponding overall air change rates of 1.34 air changes per hour (ACH) and 5.05 ACH. The fraction of this flow that is fresh air is dependent on the air handling unit’s total air flowrate and the fraction of outside air rate which varied from approximately 72% at the low air change rate to 23% at the high air change rate. Details on the room dimensions and heating ventilation and air conditioning system are provided in Table 1.

**Table 1:** Room information for testing an modeling studies.

| Parameter | Description | Value | Units | Info. Source |
| --- | --- | --- | --- | --- |
| $A_R$ | Room floor area | 126.35 | m<sup>2</sup> | Facility Staff |
| $H_R$ | Ceiling height | 2.870 | m | Facility Staff |
| $V_R$ | Room volume | 362.64 | m<sup>3</sup> | Facility Staff |
| $N_{ACH,low}$ | Air changes per hour (all flow) | 1.344 | h<sup>-1</sup> | Measured |
| $N_{ACH,high}$ | Air changes per hour (all flow) | 5.050 | h<sup>-1</sup> | Measured |
| $\eta_{fHVAC}$ | HVAC system filtration efficiency | 0.8 | - | Estimated |
| $\dot{V}_{FA,low}$ | Fresh air flow rate, low flow | 351.1 | m<sup>3</sup>/h | Facility Staff |
| $\dot{V}_{FA,high}$ | Fresh air flow rate, high flow | 421.3 | m<sup>3</sup>/h | Facility Staff |
| $\dot{V}_{tot,low}$ | Total HVAC flow rate to room, low flow | 487.6 | m<sup>3</sup>/h | Current study |
| $\dot{V}_{tot,high}$ | Total HVAC flow rate to room, high flow | 1832 | m<sup>3</sup>/h | Current study |
| $\dot{V}_{AHU,min}$ | Total air handler minimum flow (services multiple rooms) | 28,000 | m<sup>3</sup>/h | Facility Staff |
| $\dot{V}_{AHU,max}$ | Total air handler maximum flow (services multiple rooms) | 88,900 | m<sup>3</sup>/h | Facility Staff |

Normally, this particular room is equipped to accommodate 48 students and a single instructor resulting in an occupant density of 2.58 m^2^/person; however, the room layout was modified with a nominal 2.13 m separation distance between students and a 3.05 m buffer separation between the instructor and seated students in response to COVID-19. This reconfiguration of the room reduced occupant capacity to 17 (16 students and 1 instructor) decreasing the occupant density to 7.43 m^2^/person. Figure 2 shows a plan view of the reduced occupant density classroom along with relative layout of the room’s HVAC supply and return. In the schematic each student desk would be occupied by a single student.

**Figure 2:**
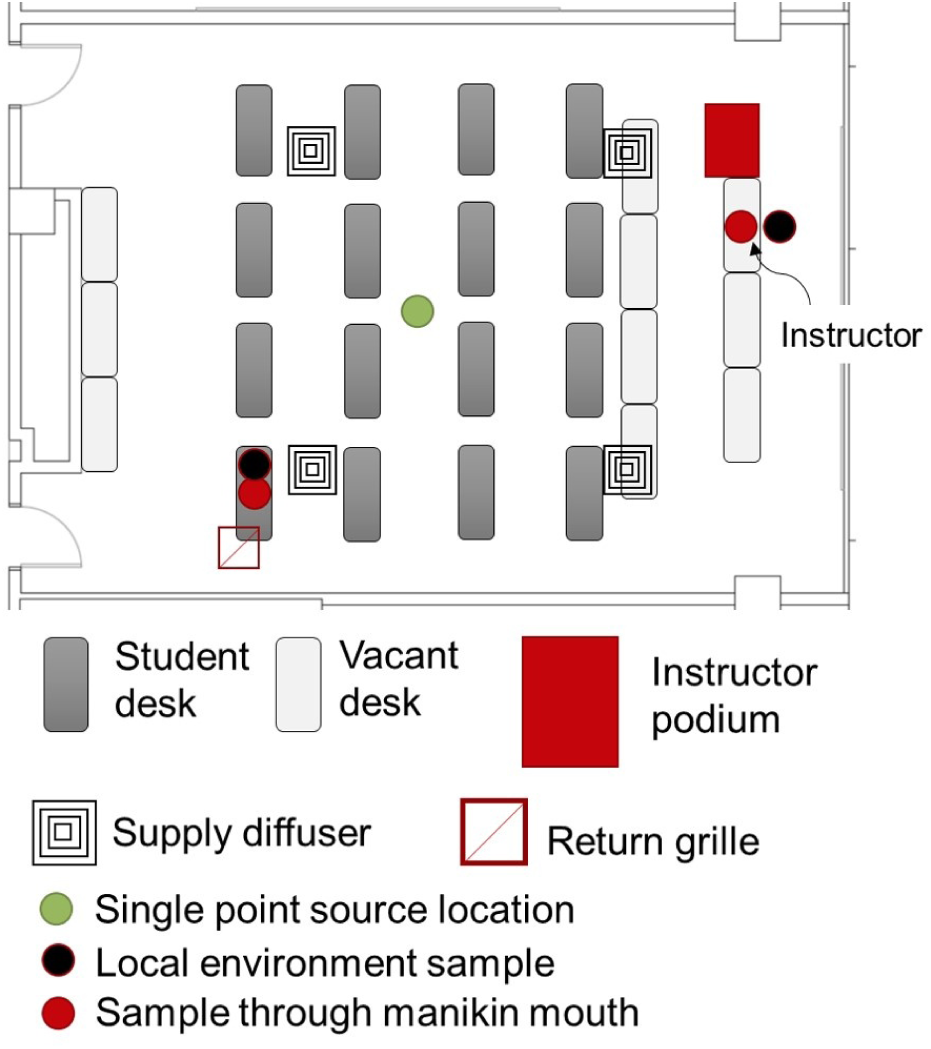
Schematic of the classroom layout used as a testbed for the current study, showing the locations of desks where student manikins were (student desk), additional desks that were empty (vacant desk), the instructor podium at the front of the room, and the supply diffusers and return.

**Figure 3:**
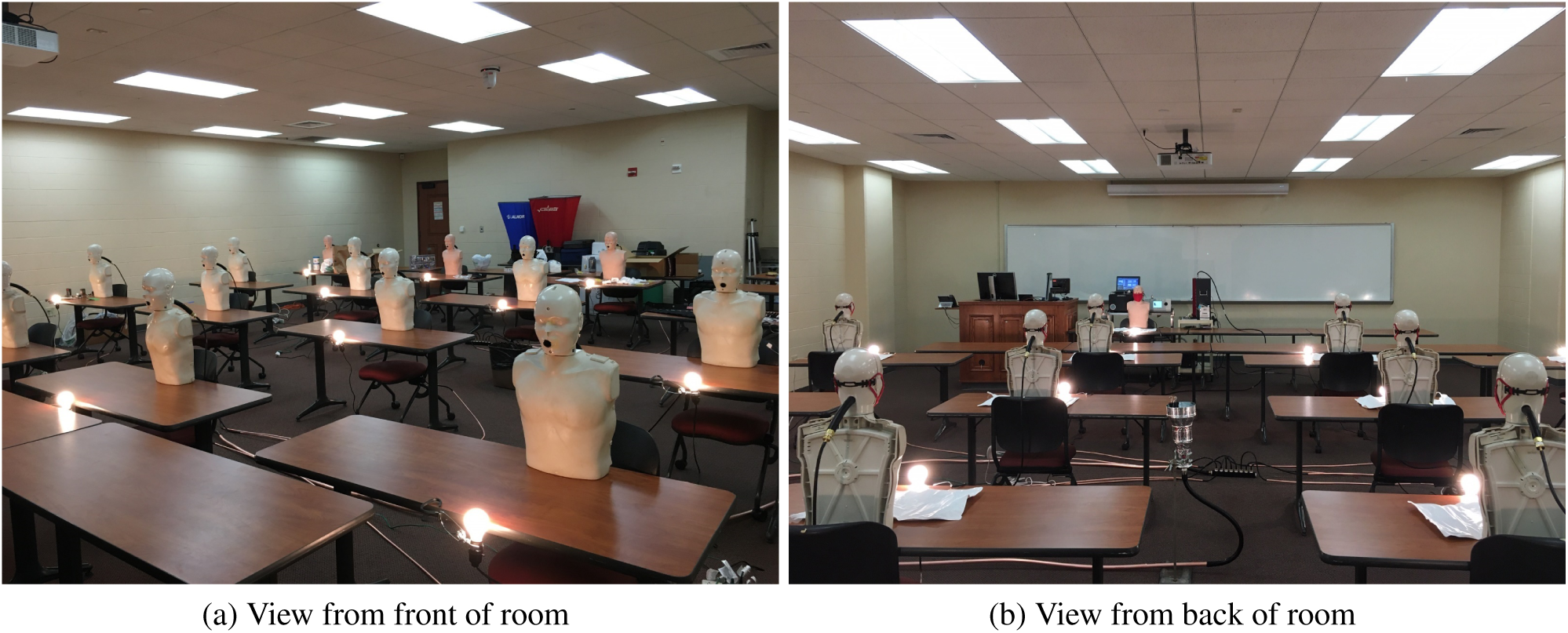
(a) Classroom space setup with manikins (a) view from the front of the room looking back and (b) from the back of the room looking forward.

To simulate occupants within the room during testing, seventeen CPR manikins were deployed and positioned at each of the sixteen student locations with the seventeenth manikin positioned at the front of the room where an instructor would be. The CPR manikins were modified to enable a 25.4-mm outside diameter (19.05-mm inside diameter) piece of conductive silicon tubing to be passed through the back of the manikin heads with the exit of the tube filling the manikins’ open mouths. This tube was connected to either the aerosol sampling instruments used to sample aerosol through the manikin’s mouth or to an aerosol source via a reducing fitting and 6.35-mm inside diameter grounded conductive silicone tubing. The ability to use the manikins for “exhaling” or “inhaling” aerosol was a key feature that enabled configuring the manikins to serve as aerosol sources to study aerosol distribution in the room to simulate an infected student or to draw air samples for measuring the effective filtration efficiency of masks installed on a manikin. Each student location also had an incandescent (75 W) lightbulb co-located to simulate sensible heat load from classroom occupants. Light bulbs were turned on during all testing performed for the work.

### 3.2 Aerosol generation and measurement

Aerosol testing was conducted in the classroom to: (1) assess validity of the well-mixed assumption in the Wells-Riley model, (2) evaluate the rate of concentration build-up/decay and loss rates of particles, and (3) obtain data on the effective filtration efficiency of various masks as worn by occupants.

A polydisperse NaCl (salt) aerosol was generated by atomizing a 20% by weight solution of NaCl and distilled water using an aerosol generator (TSI 3076) with a nominal air flow rate of 3*±*0.5 SLPM. The output of the aerosol generator was dried using a diffusion dryer (TOPAS DDU 570/H) followed in series by two aerosol neutralizers (both TSI 3077s). Following aerosol drying and charge neutralization, an axial diluter was used to reduce particle concentration and to achieve the desired total aerosol flowrate. Use of a charge neutralized aerosol provides a more conservative estimate of mask filtration efficiency relative to the use of an unneutralized aerosol. Total aerosol flowrate was varied based on the number of manikins (or sources) setup to emit aerosol to the room.

Three different types of aerosol measurement instruments were used to measure size-resolved aerosol concentrations at discrete locations within the classroom and through the mouths of the manikins. A summary of the aerosol measurement instruments is given in Table 2. The electrical low-pressure impactor (ELPI) and aerodynamic particle sizer (APS) both classify particle size based on a particle’s aerodynamic diameter, making the measurements directly comparable with regards to size. The optical particle sizer (OPS), on the other hand, sizes particles individually based on light scattering and the sizes are not directly comparable to the other two instruments without calibration. Because of this, for size-resolved information the ELPI and APS results were typically used.

**Table 2:** Aerosol size and concentration measurement instruments used for classroom measurements.

| Model | Type | Diameter Range [$\mu\text{m}$] | # Diameter Channels | Sample Flow [L/min] |
| --- | --- | --- | --- | --- |
| Dekati ELPI | Electrical low-pressure impactor (ELPI) | 0.030 – 10 | 12 | 10 |
| TSI 3321 | Aerodynamic particle sizer (APS) | 0.5-20 | 52 | 1 |
| TSI 3330 (Qty. 2) | Optical particle sizer (OPS) | 0.3-10 | 16 | 1 |

A representative NaCl particle size distribution, generated with a dilution air flow rate of 120±12 SLPM and measured by the ELPI, is shown in Figure 4. The distribution has a count median diameter (CMD) of 0.25 *µ*m, a geometric mean diameter (GMD) of 0.34 *µ*m, and a geometric standard deviation (GSD) of 1.86. Particle sizes with significant concentrations range from 0.043 *µ*m to approximately 3.2 *µ*m in aerodynamic diameter. The size distribution is not perfectly log-normal and appears to have two modes (one centered around 0.25 *µ*m and a second around 0.8 *µ*m) as indicated by the bump on the right-hand side of the distribution around 0.8 *µ*m.

**Figure 4:**
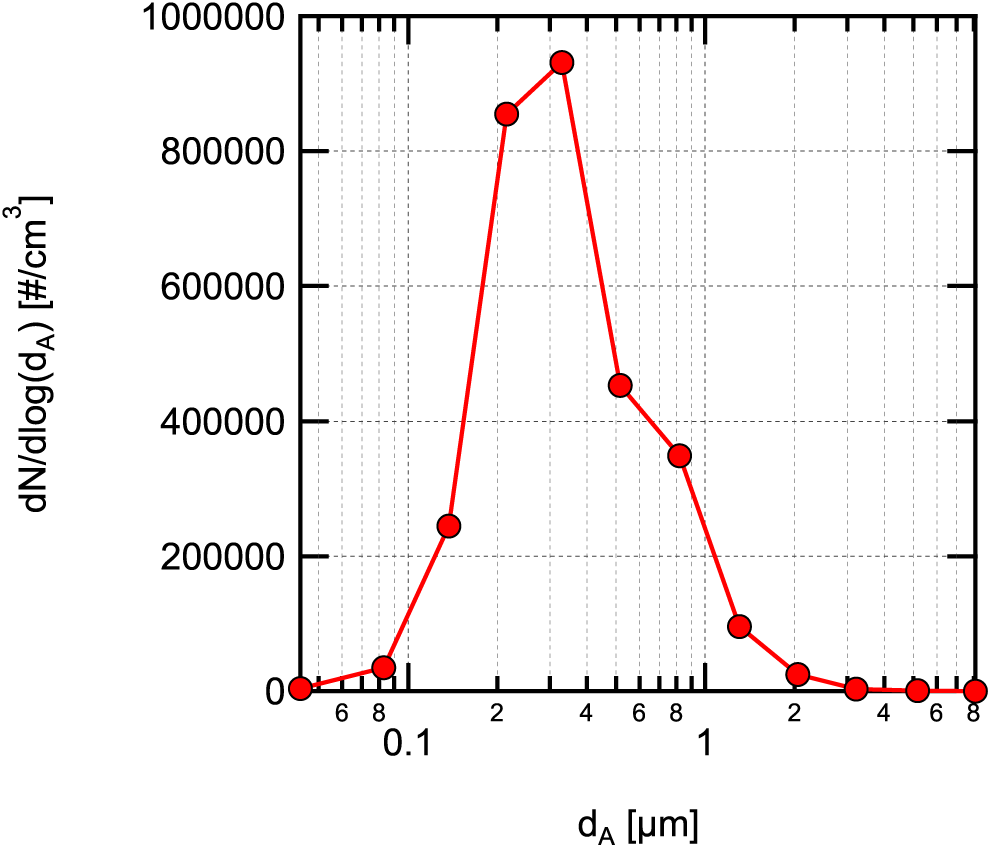
NaCl aerosol size distribution as a function of aerodynamic diameter (*d_A_*) generated by the aerosol generator measured directly after dilution.

Particles generated by breathing, speaking, and coughing, *<* 10 *µ*m in size, are typically distributed in one or two approximately log-normal modes with equilibrated count median aerodynamic diameters around 0.8 to 1.2 *µ*m and GSDs of approximately 1.3 and 1.7, respectively [15, 23]. Equilibrated particle sizes (size after initial evaporation to equilibrate with the room relative humidity) for these modes with significant concentrations range from approximately 0.2 *µ*m to 4 *µ*m in aerodynamic diameter. Therefore, the range of particle sizes tested in the current work is representative of the typical size range of aerosol particles emitted while breathing, speaking, and coughing.

The concentration of the NaCl aerosol used in the measurements performed here is several orders of magnitude higher than emitted during breathing, speaking, or coughing. A much higher concentration was used to enable discrimination of the NaCl aerosol relative to the background aerosol concentrations in the room used for the measurements. Typical background particle concentrations in the size range for the measurements were 500*±*200 cm*^−^*^3^ as measured by the ELPI for the entire size range from 43 nm to 10 *µ*m. For the NaCl aerosol emission rate used in the experiments, at the typical HVAC conditions (1.34 ACH), the steady-state concentration of NaCl aerosol was approximately 10^4^ cm*^−^*^3^, providing good discrimination relative to the background aerosol concentration.

### 3.3 Aerosol dynamics and distribution

#### 3.3.1 Aerosol dynamics

For the first set of experiments used to study the room aerosol dynamics, aerosol was dispersed from a point at the center of the room, exiting upward from a 152-mm diameter duct at an exit height of 0.97 m with an exit velocity of 6.15 cm/s giving a jet exit Reynolds number of 576. Aerosol measurements were made in the front of the room near the instructor position indicated in Figure 2 at a sampling height of 1.2 m (the approximate height of the instructor manikin’s mouth). Sampling was performed with the APS and ELPI at this location directly through the vertically oriented instrument inlets (sample travels downward into the instrument through the inlet). Sampling in the rear of the room was performed at a location near the overhead air return grille. The two OPS instruments were used at this location sampling at a height of 1.2 m through their vertically oriented instrument inlets, similar to the ELPI and APS at the front of the room. Sampling with two instruments at each location enabled a consistency check between the instruments at the same location to verify that the instruments were functioning properly. A balometer (TSI Alnor) was used to measure return and supply air flowrates at the start and end of each test.

A second experiment was conducted where the NaCl aerosol was dispersed through the mouths of all non-sampling manikins (15 total). The ELPI was setup to sample aerosol through the mouth of a manikin at the front of the room at the instructor position, and OPS2 was setup to sample through the mouth of a manikin in the back corner of the room by the overhead air return grille, see Figure 2 for locations.

A third experiment evaluated the steady-state room aerosol concentrations at lower and higher room supply air exchange rates. Aerosol was seeded steadily into the room for a duration greater than 4 times the time constant for the low room air HVAC flow of 1.34 ACH (*τ* = 1*/λ*) bringing the room aerosol concentration to steady-state as measured by the ELPI. Once steady-state was reached, the air flow supplied to the room by the HVAC system was increased to 5.05 ACH by biasing the room thermostat using a local heat source and the time evolution of the decrease in aerosol concentration was measured.

#### 3.3.2 Aerosol spatial distribution

Measurements of the aerosol spatial distribution in the room were acquired to assess the relative validity of the well-mixed room assumption in the Wells-Riley model. For these measurements, aerosol was seeded steadily into the room for a duration greater than 4 times the time constant for buildup of aerosol concentration in the room, resulting in an approximately steady-state aerosol distribution in the room. Size-resolved aerosol concentration measurements were made at discrete locations adjacent to each student position and at probable instructor locations using OPS1.

Referring to Figure 5, 4-minute time-averaged aerosol concentration measurements were made at a height of 0.97 m (3.2 ft) above the floor at positions numbered 1-17. For positions 18-24, the measurement height was increased to 1.2 m above the floor, close to the height of the mouth of the instructor manikin. OPS2 continuously measured the concentration at position 1 throughout the measurement duration. This measurement was used to adjust the OPS1 measurements for slow variations in the average room concentration during the approximately 2-hour long period required for measurements. Flow conditions for the test are summarized in Table 3.

**Figure 5:**
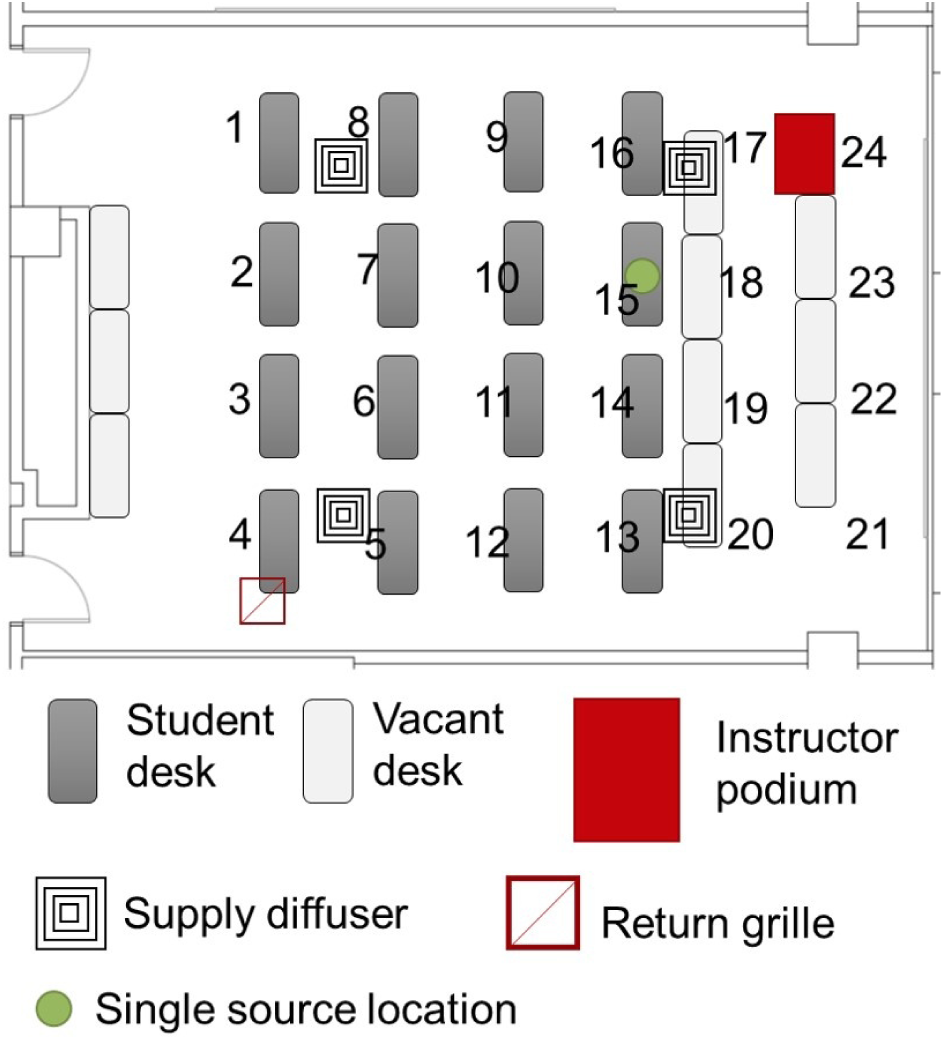
Classroom layout showing setup for mapping distribution of concentration within the classroom. Number locations indicate positions where aerosol concentration was measured. Location of single student manikin emitting aerosol during the measurements is indicated by a green dot.

**Table 3:**
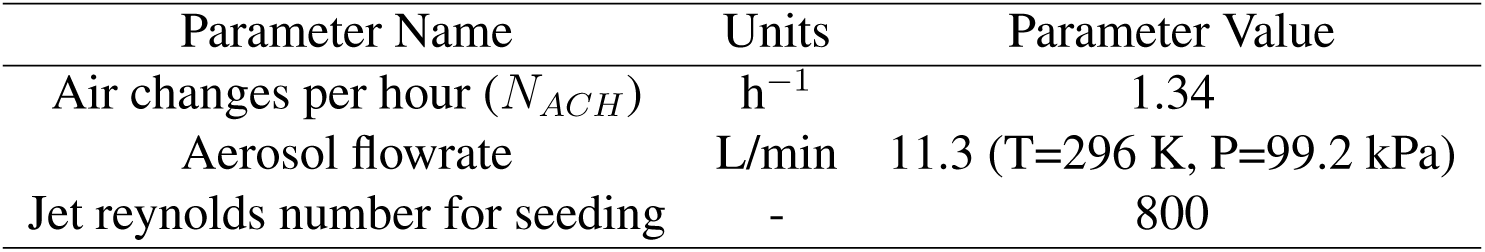
Conditions for spatially-resolved aerosol concentration measurements.

| Parameter Name | Units | Parameter Value |
| --- | --- | --- |
| Air changes per hour ($N_{ACH}$) | $\text{h}^{-1}$ | 1.34 |
| Aerosol flowrate | L/min | 11.3 (T=296 K, P=99.2 kPa) |
| Jet Reynolds number for seeding | - | 800 |

### 3.4 Effective filtration efficiency

Many locales have established requirements for wearing “face coverings” (i.e., masks) while in buildings and, in some cases, while outdoors in public spaces. These emergency orders do not generally provide detailed specifications on the face covering to be worn. As an example, the Wisconsin statewide emergency order requiring face coverings (Emergency Order #1) defines a face covering as [55]: ““Face covering” means a piece of cloth or other material that is worn to cover the nose and mouth completely. A “face covering” includes but is not limited to a bandana, a cloth face mask, a disposable or paper mask, a neck gaiter, or a religious face covering. A “face covering” does not include face shields, mesh masks, masks with holes or openings, or masks with vents.” Due to the loose definition of a face covering, these requirements result in individuals utilizing a range of means for complying such as bandanas, cloth gaiters, cloth masks of various designs, and disposable procedure masks, among others. Additionally, emergency orders requiring masks have no specifications for mask fit beyond covering “the nose and mouth completely”.

Here we evaluate various types of masks that are designed to cover the nose and mouth with the intent of providing respiratory particle protection for the wearer and those in physical proximity to the wearer. The masks tested include a commercial 4-ply knit cotton mask, a 3-ply spunbond polypropylene (material ADO Product Pro Pac Insulation Fabric) mask designed by the UW-Madison emergency operations committee (EOC) and produced by a local custom sewing manufacturer for UW-Madison (Laacke & Joys Design & Manufacturing) referred to as the EOC mask throughout, a 3-ply disposable non-medical mask with a melt-blown polypropylene center ply (Hodo single use mask) referred to as a procedure mask throughout, and an ASTM F2100 [56] level-2 rated medical surgical mask (Medicom SafeMask FreeFlow). Images of the masks as installed on the manikins for filtration measurements are shown in Figure 6. Although not an exhaustive list, these four mask are representative of a relatively broad range of filtration performance that individuals may have access to.

**Figure 6:**
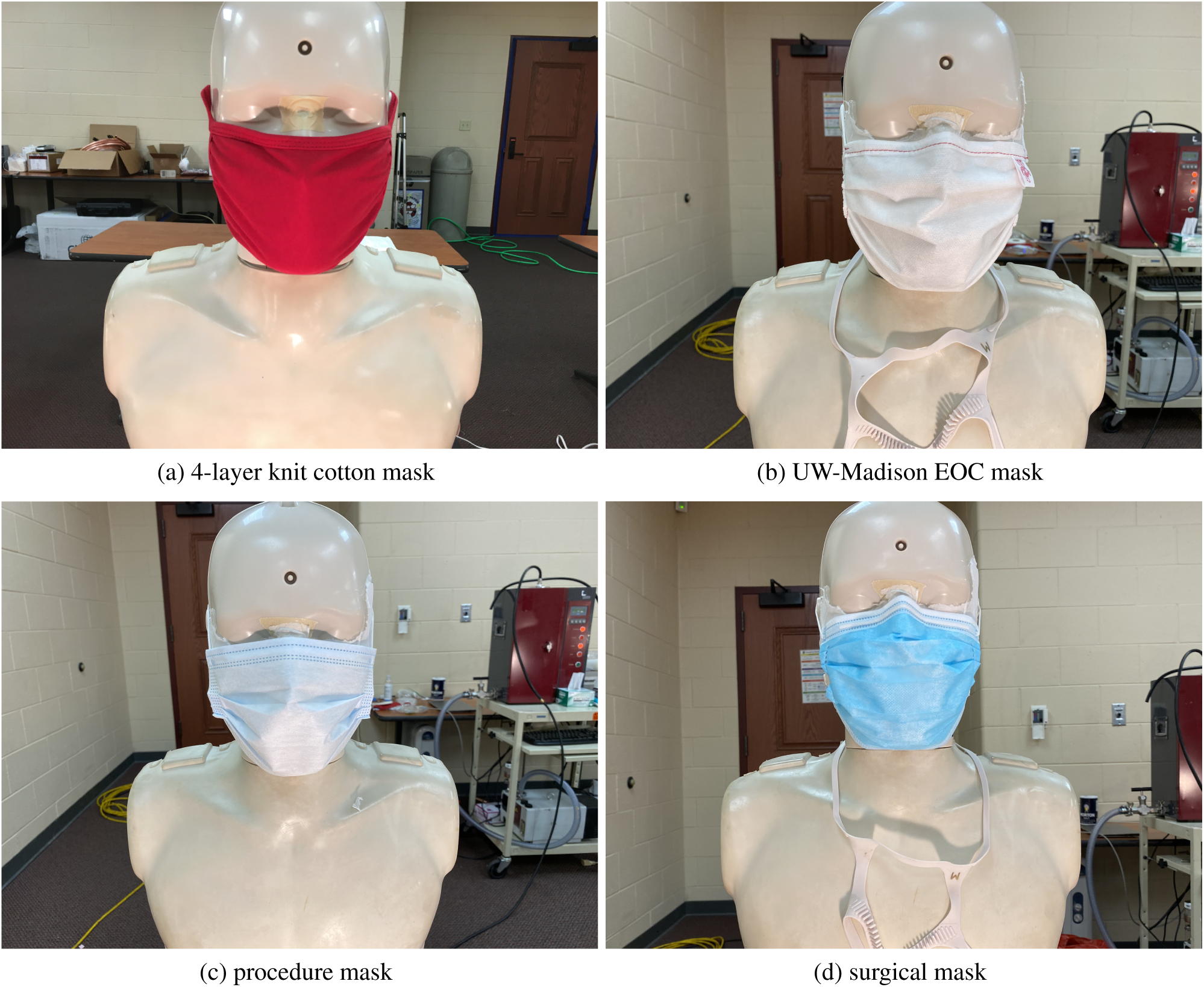
Images of masks installed on manikin for filtration testing (a) 4-layer knit cotton mask, (b) EOC mask, (c) procedure mask, and (d) surgical mask

The surface of the hard plastic CPR manikin’s face was modified to be slightly compliant to better represent a human’s face. This was done by covering the surface of the manikins face where the mask sits with a rubberized double-sided tape (Alien tape) approximately 2-mm thick. The exposed side of the double-sided tape was covered with low-lint specialty task wipes (Kimberly-Clark Kimtech Science Kimwipes). This gave a slightly compliant smooth surface that better mimics a human’s skin than the hard plastic manikin face, although the surface was still less compliant than a human’s face.

Measurements were performed with masks fit to the manikins in a way that is representative of someone intent on effective use of the mask, i.e., covering the nose and mouth completely with the formable nose piece (if present) shaped to the manikin’s face. Additionally, due to the relatively small size of the manikin’s head an adjustable mask ear saver (Seljan Company, https://earsaver.net/) was used to allow the masks to be pulled tight to the manikin’s face. The fit of the masks is close to the best achievable with each individual mask on its own. It should be noted that the 4-ply knit cotton mask did not have a formable nose piece and that the nose piece for the procedure mask did not maintain its shape well after forming. Even with trying to fit the masks well, some gaps near the nose bridge and on the sides and bottom of the mask were visible resulting in leakage around the mask. This was particularly true for the knit cotton mask and the procedure mask, where, as seen in Figures 6a and 6c, there is a clear gap between on either side of the bridge of the nose.

In a second round of testing, we evaluated the use of a mask fitter developed in collaboration with Lennon Rodgers at the UW-Madison Makerspace (Badger Seal, https://making.engr.wisc.edu/mask-fitter/), and a commercial mask fitter (Fix the Mask (FTM) mask brace, https://www.fixthemask.com/). The mask fitters are designed to seal a mask to the user’s face to minimize leakage around the mask; this enables the full filtration potential of the mask to be utilized. It should be noted that use of mask fitters requires that the masks used have reasonably low pressure drop at the flowrates typical for breathing. All of the mask materials used here fulfill that requirement with typical pressure drops (as measured in separate experiments with a 25.4 mm diameter flow area) *<* 40 Pa for a face velocity of 3.5*±*0.5 cm/s equivalent to a full mask flowrate of 28.3 L/min. Images of the EOC mask with the Badger Seal mask fitter and with the FTM mask brace installed are shown in Figure 7.

**Figure 7:**
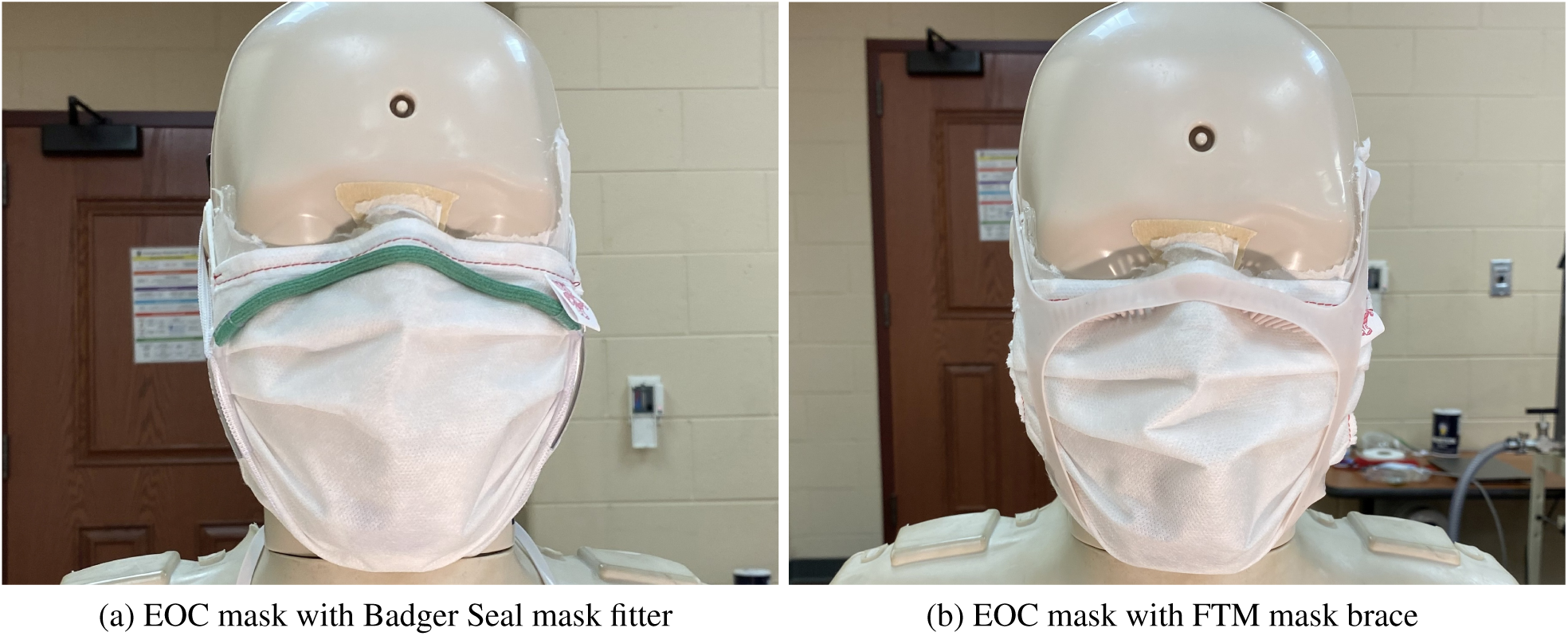
Images of EOC mask on the manikin during filtration testing with (a) badger seal mask fitter and (b) FTM mask brace installed.

Strong evidence suggests that the effective filtration efficiency of masks installed on a user’s face is often far below the filtration efficiency of the materials the mask is composed of [42,43,57]. This is illustrated by the visualization we performed with a mannequin modified to exhale a fog aerosol shown in Figure 8 wearing the procedure mask. In the image without the mask fitter installed (Figure 8a), significant leakage of fog out of the top of the mask and at the sides of the mask is seen. The image shown in Figure 8b shows the same mannequin exhalation process with the mask fitter installed. In this image, no fog is visually seen to escape. Visualization was also performed using laser Mie scattering to give higher sensitivity to low aerosol concentrations and small droplets. In that case, a small amount of aerosol was seen to flow out through the mask material. These images dramatically illustrate that inexpensive single-use masks using real filtration materials (this mask has a melt-blown polypropylene center ply) can substantially reduce aerosol emission when fit properly, but normally, significant leakage around the mask exists resulting in greatly reduced effective filtration efficiency.

**Figure 8:**
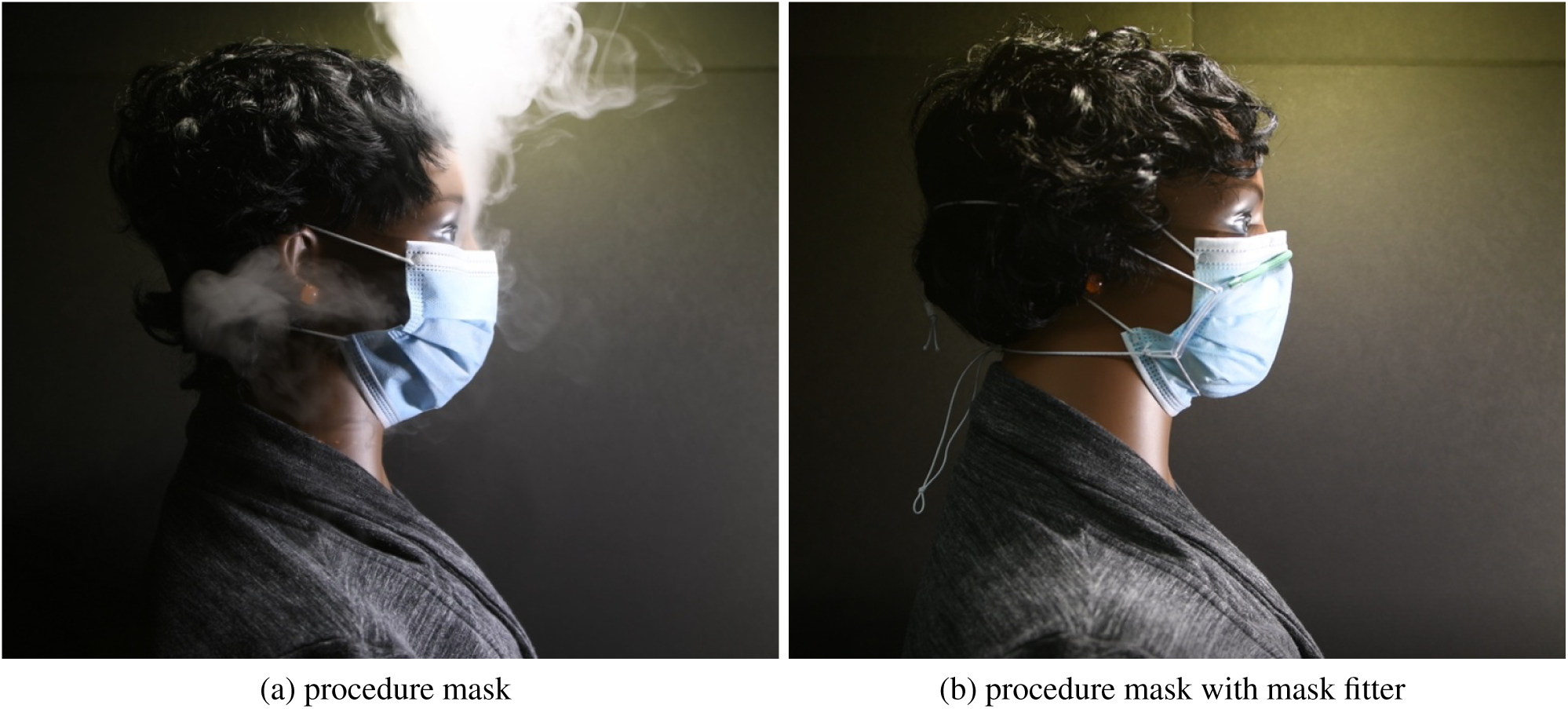
Visualization illustrating aerosol leakage for the procedure mask (a) mask as typically worn (b) mask with Badger Seal mask fitter installed.

Mask aerosol leakage makes estimating infection probabilities using the Wells-Riley model highly uncertain when trying to take into account the effective mask filtration efficiency. We define effective filtration efficiency as the filtration efficiency of the mask as worn by the user. This is lower than the mask material filtration efficiency which only considers the flow going through the mask material. In reality, given the pressure drop across the mask filtration material it is expected that in normal mask wear a large fraction of the flow will go out the sides of the mask, significantly decreasing the filtration efficiency as worn, i.e., the effective mask filtration efficiency. For example, a simple estimate of the leakage velocity for a relatively low mask pressure drop of 20 Pa using the Bernoulli equation (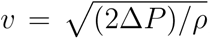) gives a potential leakage velocity of *∼* 6 m/s, which for a 1 cm^2^ leakage area, would result in a leakage flow rate of 35 L/min (assuming a constant pressure drop). This flowrate is larger than the typical breathing flowrates associated with most activities [58], indicating that if such a leakage path exists, most of the flow would follow that path and not go through the mask.

Effective filtration efficiency measurements were made by seeding the room with the same polydisperse neutralized NaCl aerosol used for aerosol dynamics and distribution measurements. Room air was continuously seeded with aerosol during measurements. Measurements were not performed until the room aerosol concentration reached an approximate steady state (*>* 3 time constants after initiating seeding at 1.34 ACH). The aerosol particle size distribution in the room measured by the ELPI after steady state had been reached in the room is shown in Figure 9. The distribution shown here was measured in the back of the room near the return grille (position 4 in Figure 5), whereas the distribution shown previously in Figure 4 was the distribution supplied to the room.

**Figure 9:**
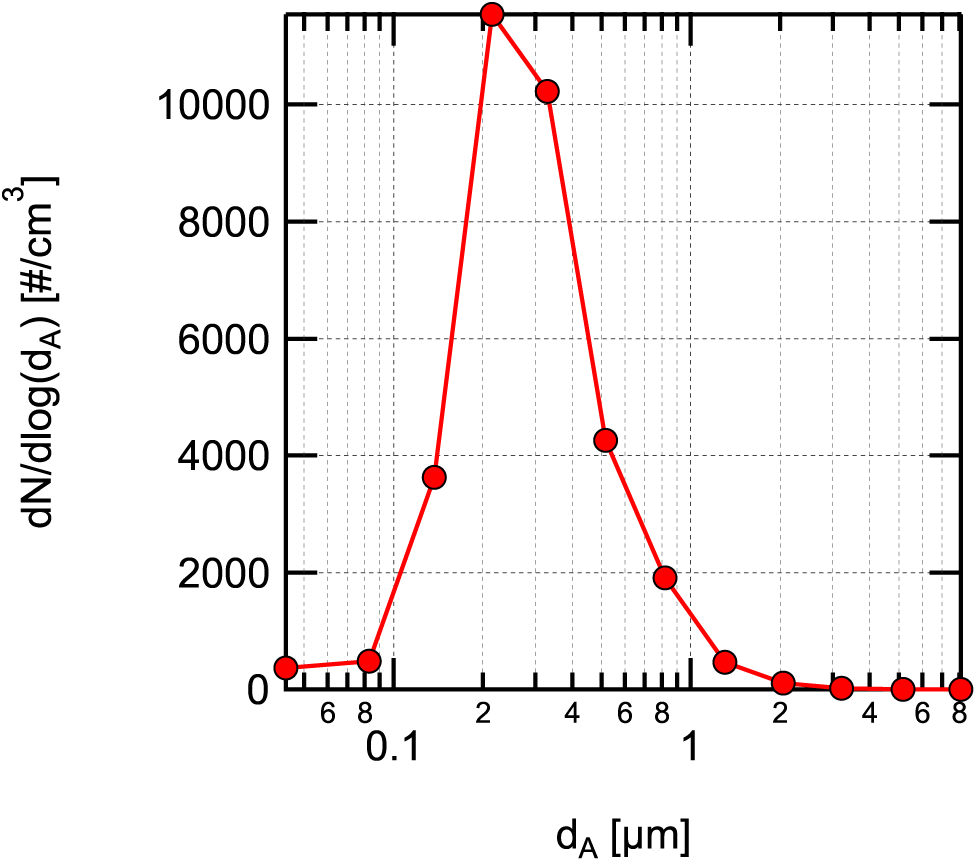
Aerosol distribution measured with the ELPI during filtration efficiency measurements measured at position 4 in Figure 5 after steady-state was reached. Error bands representing uncertainty due to random variations during the averaging interval are smaller than the symbols used and are not visible in the plot.

Effective filtration efficiency for inhalation was tested by sampling air from the room through the 19.05-mm inside diameter conductive silicone tube installed in the manikin’s mouth connected via a brass hose fitting reducer to a 6.35-mm inside diameter conductive silicon tube connected to the inlet of the ELPI. The entire sample was sent to the ELPI with a sample flowrate of 9.7 L/min. This flowrate rate is similar to inhalation rates for sitting and/or standing for adults [58]. All measurements were performed with the ELPI using the followoing procedure:

1. sample room air for 4 minutes without a mask,
2. install mask on the manikin,
3. sample room air through mask on the manikin for 4 minutes after flow equilibrates (*∼*1 min),
4. remove mask from manikin, and
5. sample room air for 4 minutes without a mask.

Measurements performed without the mask installed before and after the mask measurement were averaged when determining filtration efficiency. For the size-resolved measurement, filtration efficiency for each particle size is given by

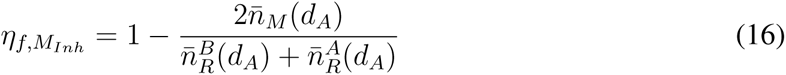

Where 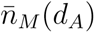 is the average number density of particles at aerodynamic diameter (*d_A_*) measured with the mask on the manikin, and 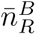 and 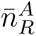 are the average number densities measured before and after with the mask off (representing the room concentration). Uncertainty in the filtration efficiency determined using Equation 16 was estimated using first-order uncertainty propagation. Uncertainty estimates include contributions due to random measurement variation during the averaging period, bias due to drift in the room concentrations, and bias estimates based on potential zero drift for each channel on the ELPI assuming a maximum zero current drift of *±*2.5 fA (see the Dekati ELPI manual for converting this to a number density for each size bin).

Total filtration efficiency based on the particle size distribution used in the measurements, shown in Figure 9, was calculated using the same equation as the size-resolved measurements, but with the size-resolved number densities replaced with the total number densities. Uncertainty was calculated in the same fashion as the size-resolved measurements just described for the total filtration efficiency. The only difference being that instead of using an assumed current drift for the zero bias, the zero bias was assumed to have a maximum number density value 150 cm*^−^*^3^ based on observations during the measurement campaign.

## 4 Results and discussion

### 4.1 Aerosol dynamics and distribution

#### 4.1.1 Aerosol dynamics

The transient response of the classroom aerosol concentration during buildup and a subsequent decay at the two measurement locations is shown in Figure 10. Data collection started 5 minutes before the initiation of aerosol seeding at 10:20 am from the single centrally-located seeding position (shown in Figure 2). Aerosol was seeded at an approximately constant rate for 1 hour after which the aerosol flow was stopped. During the period where data were collected, the supply air flow to the classroom averaged 488 m^3^/hr (287 cfm or 1.34 ACH). After the aerosol flow was stopped at 11:20 am, the air exchange in the room and settling reduced the aerosol concentration in the room.

**Figure 10:**
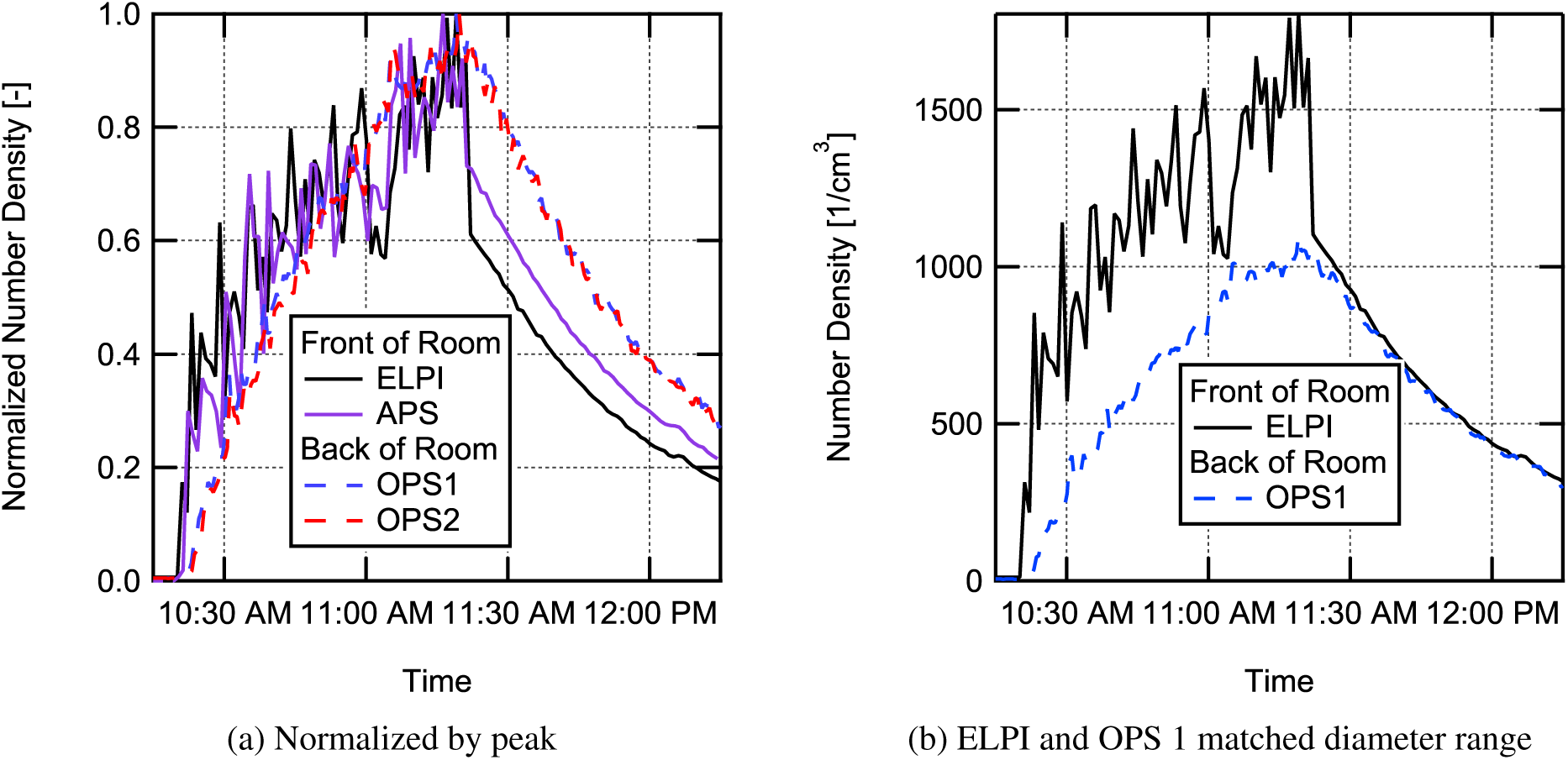
Transient response of aerosol concentration in the classroom during buildup of aerosol from a single source located near the center of the room and during the decay of aerosol following shutoff of the aerosol source. (a) Aerosol concentrations normalized by their peak value for all four instruments used and (b) measured time-resolved aerosol concentrations in the front and rear of the room for a similar range of diameters. Aerosol seeding was started at 10:20 am and stopped at 11:20 am.

Results are shown in Figure 10a as normalized concentration. This was done due to the differences in diameter resolution for the different instruments. This prevented total concentrations for the exact same size range from being calculated for all the three instrument types. However, the size range for the ELPI results were adjusted to roughly match the APS and OPS measurement range and measurements from all instruments were binned to a matched 1 min per data point time resolution. This gave peak concentrations of roughly 1500 cm*^−^*^3^ (in comparison, the background concentration before aerosol seeding was 11 cm*^−^*^3^ in this size range) based on ELPI measurements in the aerodynamic diameter size range from 0.66 *µ*m to 10 *µ*m (the majority of particles are *<* 3.2 *µ*m in aerodynamic diameter). The APS results showed a slightly lower peak total concentration of approximately 1000 cm*^−^*^3^. This is likely due to the reduced counting efficiency for the APS below 0.9 *µ*m in size (the counting efficiency at 0.5 *µ*m has been measured previously to be 30% increasing to a value of 100% at 0.9 *µ*m) [59]. The OPS instruments both measured peak total concentrations of approximately 1100 cm*^−^*^3^ for a measurement range corresponding approximately to 0.73 *µ*m to 10 *µ*m in aerodynamic diameter.

Once the aerosol was turned on at 10:20 am, the instruments at the front of the room saw an abrupt increase in aerosol concentration followed by a somewhat noisy, approximately exponential, rise in concentration. The OPS instruments at the rear of the room near the air return showed a much smoother rise in aerosol concentration. This indicates some level of non-uniformity in the aerosol distribution within the room and that the aerosol is more uniformly mixed in the rear location. This is likely due to the proximity of the rear sampling location to the air return. Interestingly, once the aerosol source is turned off, the concentration decay for both instruments at the front of the room is almost completely smooth. Perhaps more interesting is that the absolute total concentrations shown in Figure 10b for the ELPI and OPS1 are very similar in magnitude for a similar size range, indicating that the distribution in the room becomes nearly uniform relatively quickly with the aerosol source turned off. Additionally, the decay at both the front and rear of the room have nearly the same time constant for the same range of particle sizes.

Overall, the results for the single source at the center of the room suggest a relatively uniform buildup of aerosol concentration in the room with some local non-uniformity associated with turbulent fluctuations from the plume (Re*_D_*=576) being emitted at the center of the room. The fact that the difference in concentration between the front and back of the classroom varied but the differences were modest helps support the well-mixed assumption for the Wells-Riley model. Additionally, the size-resolved data indicate that the loss rate for all particle sizes during the period were the aerosol was shutoff is similar and only modest size dependence was seen in loss rate for particle sizes in the range emitted during the experiments.

A second experiment was conducted where the NaCl aerosol was dispersed through the mouths of all non-sampling manikins (15 total). The ELPI was setup to sample aerosol through the mouth of a manikin at the front of the room at the instructor position, and OPS2 was setup to sample through the mouth of a manikin in the back corner of the room by the overhead air return grille, see Figure 2 for locations. The measured total concentration buildup and decay in this scenario was similar to the data for the single-source location. The primary differences were that significantly less temporal variation in total concentration was seen for the ELPI data at the front of the room, and a smaller difference was seen in total concentration between the front and rear of the room due to the increase in number of source locations.

An additional run evaluated the steady-state room aerosol concentrations at lower and higher room air exchange rates. Aerosol was seeded steadily into the room for a duration greater than 4 times the time constant for the low HVAC flow of 1.34 ACH, bringing the room aerosol concentration to steady-state as measured by the ELPI. Once a steady-state was reached, the air flow supplied to the room by the HVAC system was increased to 5.05 ACH. The factor of 3.8 increase in room air volume flow rate from 1.34 ACH to 5.05 ACH (488 m^3^/hr to 1,830 m^3^/hr) resulted in the average steady-state concentration decreasing from 5500 cm*^−^*^3^ to 2100 cm*^−^*^3^ based on ELPI measurements for the entire range of particle sizes measured at the instructor position. This decrease in aerosol concentration is shown in Figure 11.

**Figure 11:**
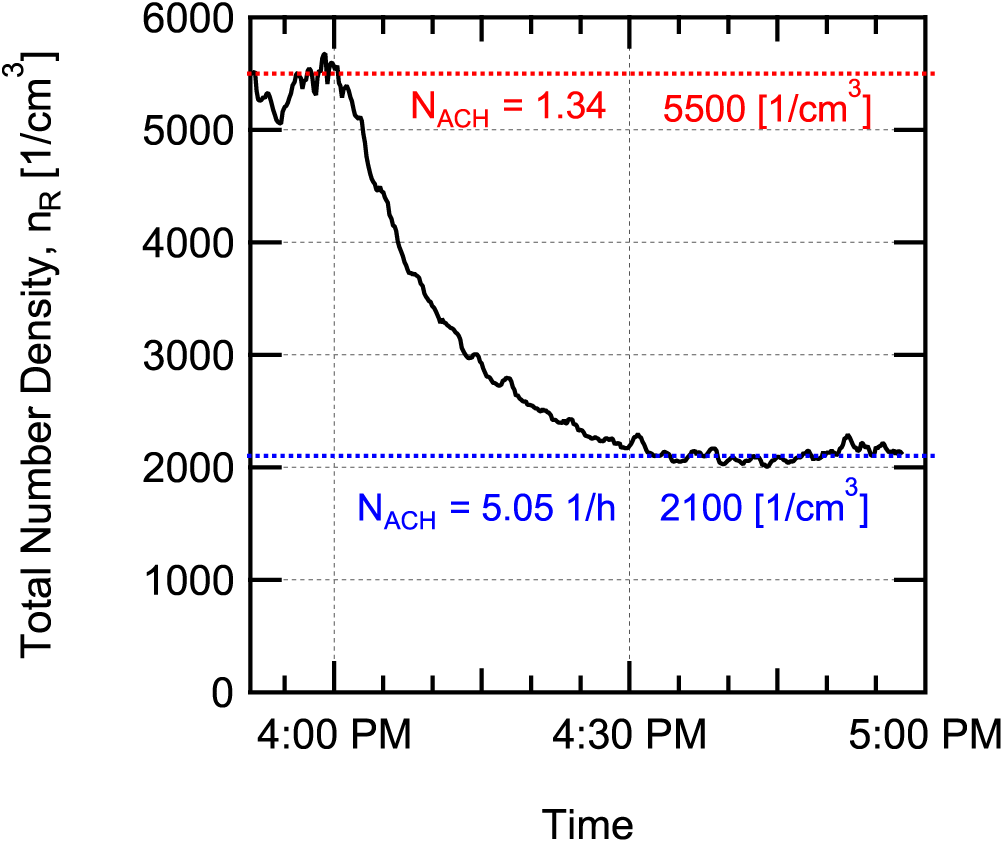
Decrease in aerosol concentration in the room when switching from low flowrate of 1.34 ACH to a high flowrate of 5.05 ACH while constantly seeding aerosol into the room.

Both the buildup of aerosol when the seeding was first started with the HVAC at 1.34 ACH, and the decrease in aerosol when the HVAC flow was increased to 5.05 ACH were curve fit to the time-dependent concentration equation for a perfectly mixed room in Equation 7 letting 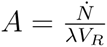. Fitting the rise in concentration for the ELPI data gave a value of *λ_low_* = 1.69 *±* 0.07 hr*^−^*^1^ for the low HVAC flowrate of 1.34 ACH and fitting the fall in concentration when switching to high HVAC flow gave a value of *λ_high_* = 5.40 *±* 0.18 hr*^−^*^1^ for the 5.05 ACH flowrate. The number of air changes per hour equals the first order loss rate due to fresh air exchange in the room. For the current work, it is assumed that the air coming in from the HVAC system was, essentially, 100% fresh air since the air handler servicing the room also services a number of other spaces and the entire building was largely unoccupied due to the ongoing pandemic.

The curve fit first-order loss coefficients are higher than that due to air exchange alone (*λ_low_* = 1.69 hr*^−^*^1^ *>* 1.34 ACH and *λ_high_* = 5.40 hr*^−^*^1^ *>* 5.05 ACH). This indicates additional loss mechanisms are present such as settling in the room or in the HVAC system. The value of total loss rate for the other mechanisms is determined by subtracting the loss rate due to air exchange from the total loss rate, giving values of *λ_other,low_* = 0.35 hr*^−^*^1^ and *λ_other,high_* = 0.35 hr*^−^*^1^ because the same value is arrived at using either the low or high flow results we assume *λ_other_* = 0.35 *±*0.1 hr*^−^*^1^, rounding the uncertainty for simplicity sake. This value corresponds to the expected loss rate for 1.2 *µ*m diameter salt particles falling from an average height of 1.07 m (average height from manikins’ mouths to a surface, accounting for tables in the room). This diameter is larger than the count median aerodynamic diameter of the particles seeded, indicating that losses in the air handling system ductwork are also possibly important, or that there is some size dependence of the loss rate (as one would expect [60]). This value will be used later in the Wells-Riley model to account for losses due to particle settling in the room and losses in the HVAC duct work.

#### 4.1.2 Aerosol spatial distribution

Figure 12 shows interpolated distribution maps derived from the discrete location concentration measurements of aerosol with a single student manikin emitting aerosol (see location in Figure 5). Results are shown for two aerodynamic diameters, 0.728 *µ*m and 2.71 *µ*m (particle size from the OPS instruments was estimated based on comparison of size distributions with the ELPI results and finding a single diameter scaling factor that gave the best agreement between the distributions). The results are normalized by the lowest concentration in the room for each diameter and case shown, indicating the relative non-uniformity in the room. Measurements were performed with no mask on the manikin (Figures 12a and 12b) and with the manikin wearing a four-layer knit cloth face mask (see Figure 6a for a picture of the mask, results are shown in Figures 12c and 12d).

**Figure 12:**
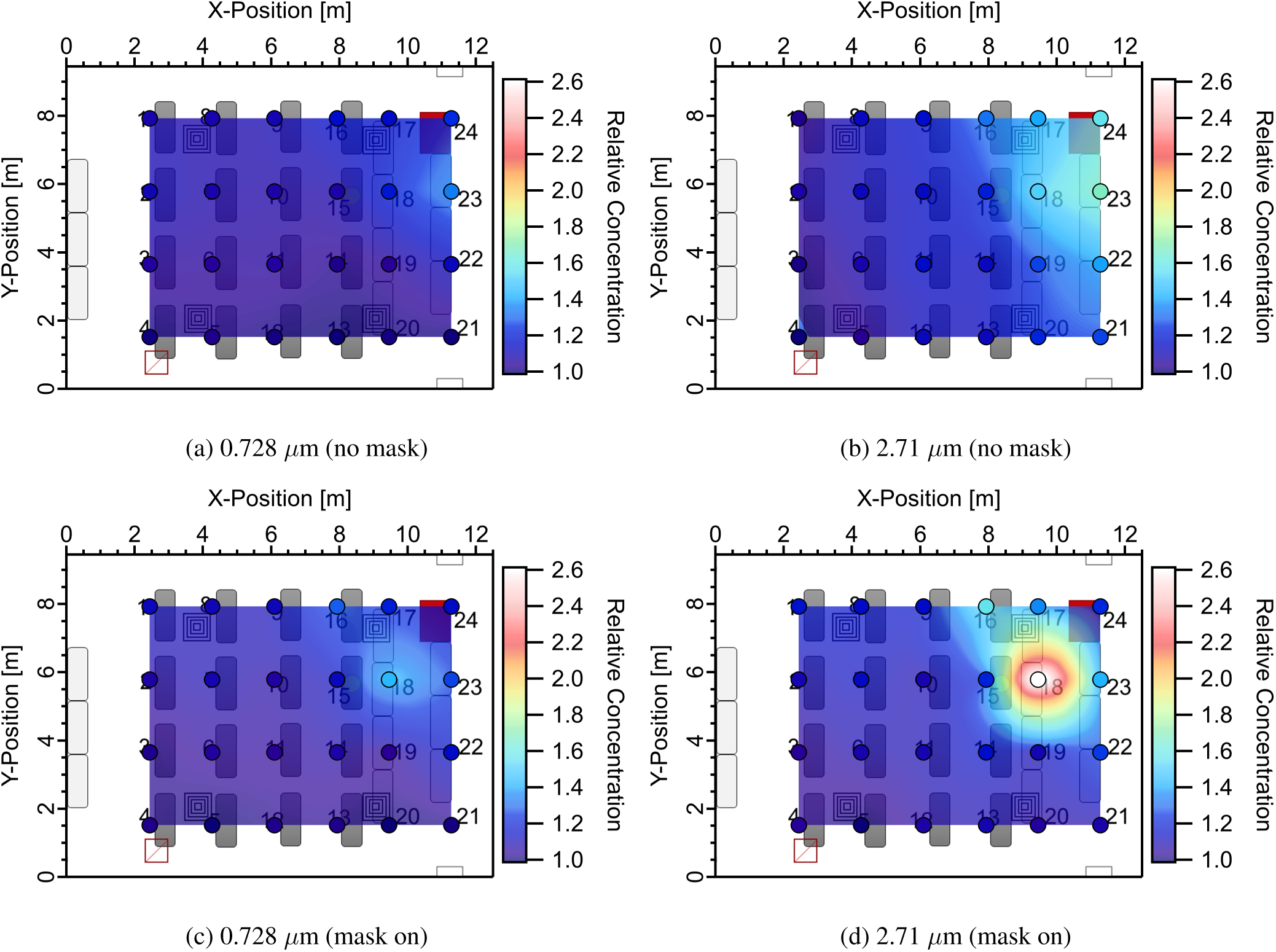
Relative concentration point measurements (markers) and spatially interpolated relative concentration distributions for (a) no masks donned and average aerosol diameter of 0.728 *µ*m and (b) 2.71 *µ*m; (c) masks on and average aerosol diameter of 0.728 *µ*m and (d) 2.71 *µ*m.

All results indicate some level of spatial non-uniformity in the aerosol distribution in the room, even under steady-state conditions. This is not at all unexpected; obviously, at the exit of the manikin’s mouth the aerosol concentration will be maximum. As the jet of aerosol emitted from the manikin’s mouth entrains air and mixes the concentration will decrease. The level of non-uniformity appears to be a function of both the presence or absence of a mask, and the size of the particles.

The presence of the mask clearly changes the penetration of aerosol in the direction that the jet exits the manikin’s mouth (towards the front of the classroom *→* increasing x-position). Without a mask, higher concentrations are seen further from the manikin location in the x-direction in Figure 12. For example, at location 23 directly in front of the emitting manikin near where an instruct might stand, with no mask on, the relative concentration of 2.71 *µ*m particles is 1.67, whereas with a mask on, the relative concentration is 1.43. The use of a mask, even the common face covering used here, appears to localize the high concentration region closer to the emitting manikin. As will be shown later, the filtration provided by the mask also reduces overall aerosol concentrations in the room. Therefore, wearing a mask has multiple benefits for reducing infection probability, by limiting the penetration of aerosol in the direction of aerosol emission and reduction of overall aerosol concentrations.

Dependence of the distributions on particle size is also seen in Figure 12. The same distribution shape is seen for both particle sizes for both cases (no mask and mask on). However, the magnitude of non-uniformity appears to be smaller for the smaller particle size (0.728 *µ*m). This suggests that there may be a significant dependence of the first order loss rate, due to settling, on particle size. The mechanism for the observed non-uniformity could be explain by a higher loss rate due to settling for larger particles. This would result in lower concentrations of large particles farther from the source and would increase the peak relative concentration for larger particles since the distributions are normalized by the smallest concentration in the room for each case and diameter. This implies that smaller particle sizes are more uniformly distributed in the room and that the well-mixed assumption holds better for these particle sizes.

In all cases shown in Figure 12, the concentration is elevated at the front of the classroom where the emitting manikin is located and decreases toward the back of the classroom. It is interesting that the position closest to the overhead return at the back of the classroom vent has the lowest concentration (within our field of view for the measurements) in the room for all cases. At first this seems somewhat counterintuitive, however, this can be explained by the fact that air from the entire room is pulled to the return vent and, as such, the emitted aerosol is diluted to the approximate average concentration in the room at this point (concentrations outside of the area measured may be lower than in the field of view).

One of the weaknesses of the current measurements is that they are relatively sparse in the x-y directions in Figure 12, and even more so in the vertical direction. The net results of this is that some sampling bias exists in the data, particularly due to the lack of vertical resolution. One can imagine at sample point 18, particularly in the case with no mask, that measurements directly along the centerline of the jet issuing from the manikin’s mouth will have much higher concentrations than locations a 1 meter above or below this vertical location. Therefore, the results need to be interpreted with this understanding in mind. As one moves further away from the source location, vertical stratification should become smaller, but may potentially still be significant. The results presented here do not provide information on the vertical stratification of aerosol in the room. However, the relatively modest non-uniformity in concentrations measured over the majority of the area of the room indicates that the well-mixed model assumption in the Wells-Riley model should result in only modest errors in the infection probabilities predicted by the model and the uncertainty this introduces is similar or smaller in magnitude than the uncertainty in some model input parameters, for example the quanta emission rate.

### 4.2 Mask effective filtration efficiency

Size-resolved inhalation effective filtration efficiencies for the four masks tested (knit cotton mask, EOC mask, procedure mask, and surgical mask) are shown in Figure 13 for the three different cases tested: without a fitter, with the Badger Seal mask fitter, and with the FTM mask fitter. Total effective filtration efficiencies for the particle size distribution used in the current measurements (shown in Figure 9) are shown in Figure 14, and the values are provided in Table 4

**Figure 13:**
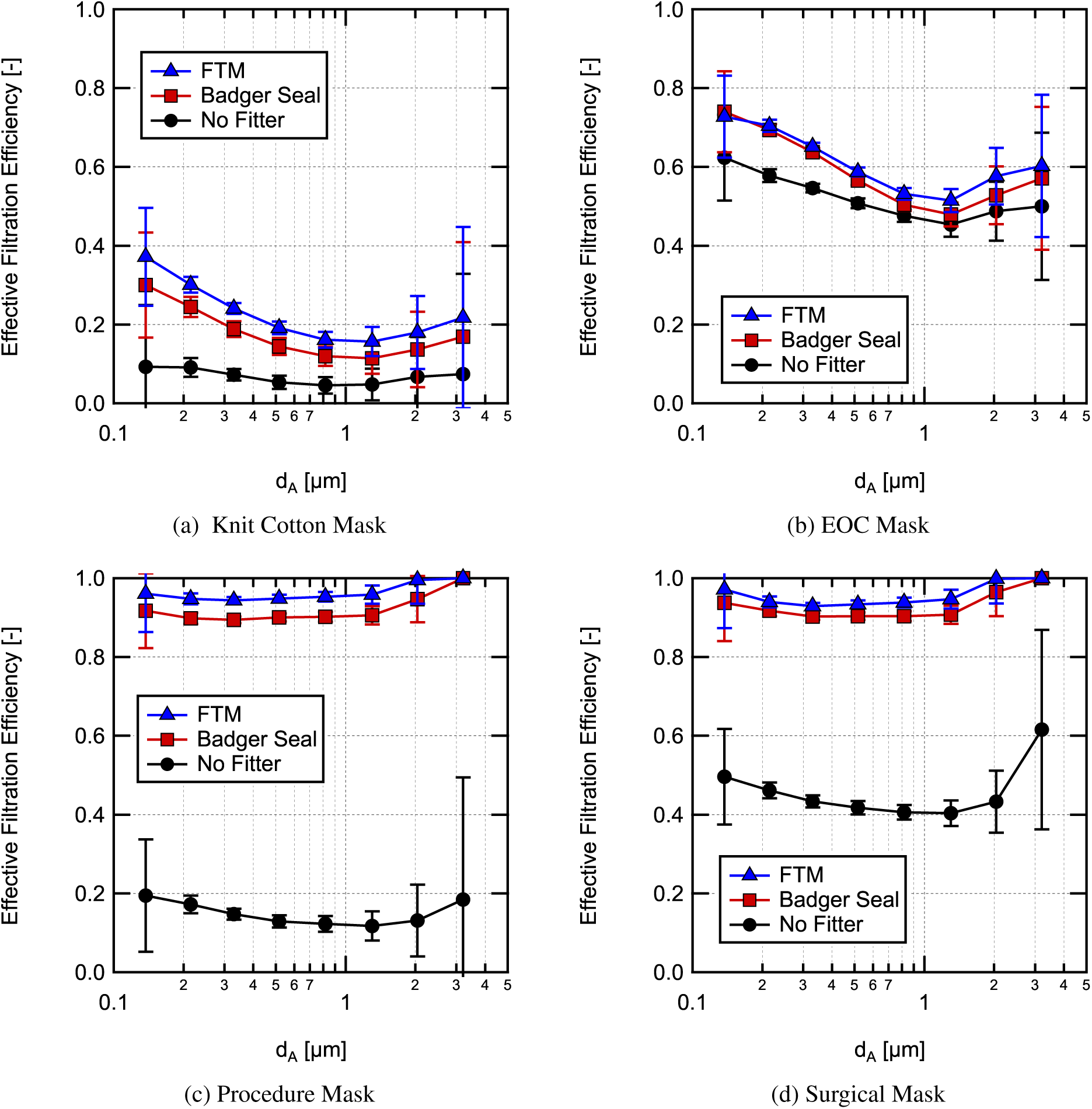
Size-resolved effective filtration efficiency for inhalation measured in a classroom without a mask fitter used, with a Badger Seal mask fitter, and with a FTM mask brace installed for (a) the 4-ply knit cotton mask, (b) the EOC 3-ply spunbond polypropylene mask, (c) a single use procedure mask, and (d) an ASTM F2100 level-2 rated surgical mask.

**Figure 14:**
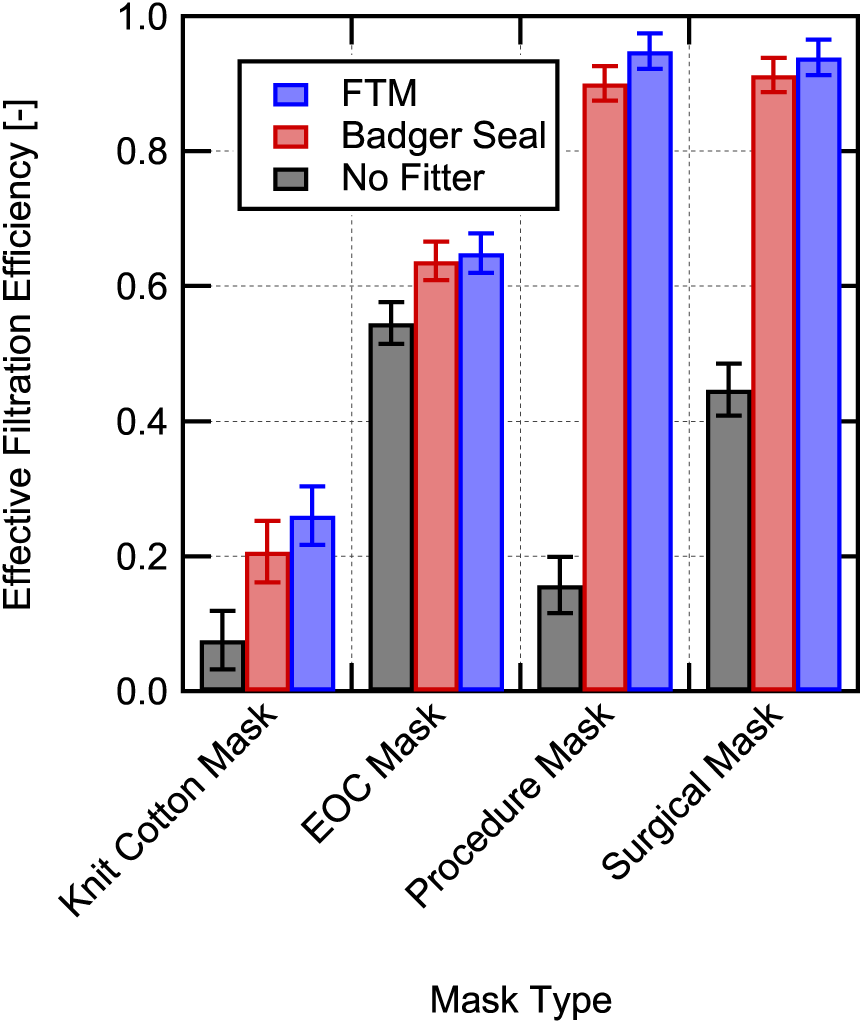
Overall effective filtration efficiency for inhalation measured using ther aerosol size distribution shown in Figure 9 for the four masks tested in this work measured without a mask fitter, with the Badger Seal mask fitter used, and the FTM mask brace used.

**Table 4:** Measured total effective filtration efficiencies and associated uncertainties during simulated inhalation for masks installed on a manikin in a classroom setting. Estimated fraction of air leakage (*f_L_*) and its associated uncertainty are also provided based on Equation 17.

| Abbrev. | Mask | Fitter | $\eta_{f,M_{inh}}$ | $\eta_{f,M_{inh}}$ Uncertainty | $f_L$ | $f_L$ Uncertainty |
| --- | --- | --- | --- | --- | --- | --- |
| KCM | Knit Cotton Mask | None | 0.075 | $\pm 0.043$ | 0.710 | $\pm 0.661$ |
| KCM-B | Knit Cotton Mask | Badger Seal | 0.206 | $\pm 0.045$ | - | - |
| KCM-F | Knit Cotton Mask | FTM | 0.260 | $\pm 0.043$ | - | - |
| EOCM | EOC Mask | None | 0.545 | $\pm 0.031$ | 0.160 | $\pm 0.084$ |
| EOCM-B | EOC Mask | Badger Seal | 0.637 | $\pm 0.029$ | - | - |
| EOCM-F | EOC Mask | FTM | 0.649 | $\pm 0.029$ | - | - |
| PM | Procedure Mask | None | 0.158 | $\pm 0.042$ | 0.834 | $\pm 0.053$ |
| PM-B | Procedure Mask | Badger Seal | 0.900 | $\pm 0.025$ | - | - |
| PM-F | Procedure Mask | FTM | 0.949 | $\pm 0.026$ | - | - |
| SM | Surgical Mask | None | 0.447 | $\pm 0.038$ | 0.524 | $\pm 0.050$ |
| SM-B | Surgical Mask | Badger Seal | 0.913 | $\pm 0.025$ | - | - |
| SM-F | Surgical Mask | FTM | 0.939 | $\pm 0.026$ | - | - |

The 4-ply knit cotton cloth mask (Figure 13a) without a mask fitter used has the lowest effective filtration efficiency in the size range tested up to 3.2 *µ*m, with an overall effective filtration efficiency of 7.5*±*4.3%. Even with the use of a mask fitter or mask brace the filtration efficiency is relatively low (26.0*±* 4.3% with the FTM mask brace), but is significantly improved by greater than a factor of two over the entire size range. It should be noted that, for sizes greater than 3.2 *µ*m in aerodynamic diameter, the filtration efficiency of the mask will continue to increase due to increasing interception and impaction filtration capture modes as particle size increases [19] providing effective filtration of droplets *>* 10 *µ*m in diameter. Filtration efficiency of the mask material is a function of the face velocity. Here relatively low breathing rates were simulated, with a higher face velocity the material filtration efficiency at larger particle sizes will increase due to the velocity dependence of the impaction capture mechanism, whereas filtration at small particles sizes will decrease due to a reduction in diffusion capture. Therefore, for high velocities associated with coughing and sneezing the mask filtration efficiency for larger particle sizes *>* 2 *µ*m is expected to improve.

The EOC mask, composed of 3-layers of spunbond polypropylene (Figure 13b, had much higher filtration efficiency than the knit cotton mask. Additionally, the mask fit and sealed around the manikin’s face well as evidenced by only a moderate filtration efficiency increase when using either of the mask fitters. The overall filtration efficiency without a mask fitter was 54.5 *±*3.1% and was 64.9*±*2.9% with the FTM mask brace for the aerosol distribution used here. As with the knit cotton mask, filtration efficiency is expected to continue to increase for larger particle sizes due to increased interception and impaction capture and effective filtration efficiencies should be quite high for 10 *µ*m and larger particles.

Of all of the mask effective filtration efficiency results, those for the single-use procedure mask demonstrate the importance of mask fit on effective filtration efficiency the best. Without a mask fitter, the size-resolved efficiency is *<* 20% for the entire range measured. With a mask fitter, the size-resolved filtration efficiency is generally greater than 90% with either mask fitter and around 95% or greater with the FTM mask brace. It should be noted that the Badger Seal is able to achieve similar performance to the FTM mask brace and in a repeat data set (not shown here) the filtration efficiency using both was around 95% and the two performed the same within the measurement uncertainty. This does indicate however that there is some sensitivity to user installation of the mask fitter. Measurements were also performed with the procedure mask with and without a Badger Seal mask fitter using a fit tester (TSI PortaCount Pro+8038) and with a well and poorly adjusted mask fitter and these results showed similar trends (fit test results are provided in the supporting information for the paper).

The low overall effective filtration efficiency of 15.8*±*4.2%, shown in Figure 14, for the single-use procedure mask without a mask fitter was due to the poor natural fit of the mask when not using the mask fitter, as illustrated in the visualization in Figure 8. Using a mask fitter enables the mask to achieve filtration efficiencies close to that of the material filtration efficiency which is close to 100% for larger paricle sizes (*>* 2 *µ*m). This demonstrates that single-use masks with a meltblown center ply can achieve excellent aerosol protection for the wearer, and those around them (assuming similar performance for exhalation), when used in conjunction with a mask fitter. One note of caution is that single-use disposable masks can vary greatly in their filtration performance even when they incorporate a meltblown center ply.

The surgical mask results demonstrate that the higher quality surgical mask provides better fit to the user’s face than the inexpensive single-use procedure mask. This can be seen directly in the images shown in Figure 6 for the procedure and surgical masks where is clear the the procedure mask is unable to conform to the bridge of the manikin’s nose. Even with the better fit, the surgical mask without a fitter significantly under performs relative to the mask worn with a mask fitter. Without a mask fitter the overall filtration efficiency for the aerosol distribution used was 44.6*±*3.8% whereas with the Badger Seal fitter it was 91.3*±*2.5% and with the FTM mask brace it was 93.9*±*2.6%.

The measured filtration efficiencies with and without the mask fitters can be used to estimate the fraction of the flow leaking around the mask for the different masks by making the following assumptions: (1) filtration efficiency does not change appreciably due to the difference in flow rate through the mask and (2) the FTM mask fitter case seals the mask perfectly and there is no leakage for this case. Both of these assumptions will tend to underestimate the leakage rate so these estimates should provide a minimum estimate of the leakage rate. Using the filtration efficiencies for the cases with and without leakage one can derive the following expression for the fraction of flow leaking around the mask (fraction of air not going through the mask material)

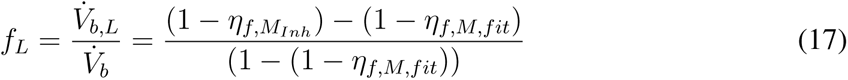

where *f_L_* is the fraction of flow leaking around the mask, *V_b,L_* is the leakage volumetric flowrate, *η_f,MInh_* is the effective filtration efficiency with leakage present, and *η_f,M,fit_* is the filtration efficiency of the mask materials (no leakage) assumed to be the same as the filtration efficiency when using the FTM mask fitter.

The estimated leakage fractions for the case with no mask fitters are shown in Figure 15 and are also provided in Table 4. The results help clarify the magnitude of leakage that is occuring around the masks. For the poorly fitting knit cotton mask and procedure mask, the leakage around the masks is estimated to be on the order of 70-85%, although the estimate for the knit cotton mask is highly uncertain due to the the low filtration efficiency of the mask. The surgical mask which visually fits much better, still has an estimated leakage of 52%. Finally, the EOC mask which fit quite well has considerably less leakage at around 15%. These estimates help emphasize the difficulty in achieving good mask fit without design features or additional devices to help seal masks.

**Figure 15:**
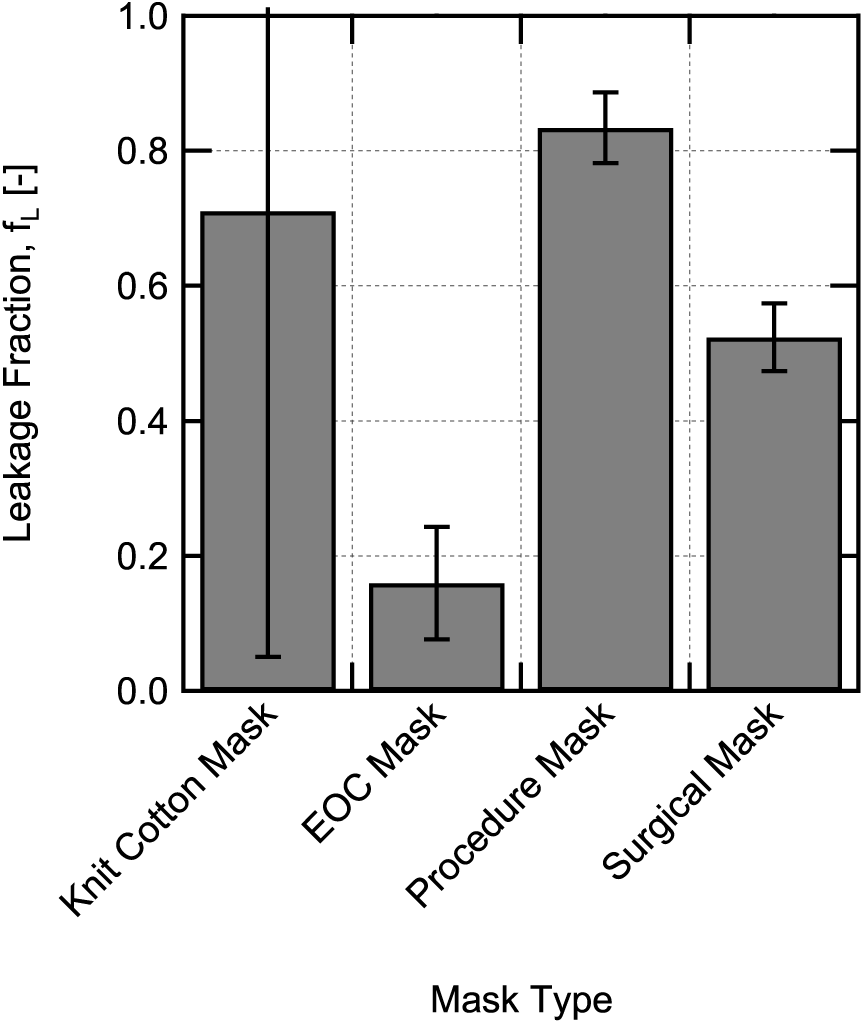
Estimated fraction of flow that leaks around the masks without use of a mask fitter, calculated using Equation 17 and the overall filtration efficiencies from Table 4.

It is interesting that with the use of a mask fitter the single use procedure mask and the the surgical mask provide effective filtration efficiencies that are the same within the uncertainty of the measurements. It is also important to note, that the washable EOC mask (this mask has been verified to maintain filtration performance at least up to 5 washes) actually out performs the surgical mask when both are used without a mask fitter. This interesting result suggests that masks made with commonly available materials like spunbond polypropylene (note that spunbond polypropylene is available as landscape fabric, insulation fabric, crop row cover material, reusable grocery bags, and is also being sold directly now precut for home mask making) can provide good effective filtration efficiencies and can even outperform ASTM level-2 rated masks in effective filtration efficiency (at least under the conditions tested here).

The filtration efficiencies for each of the mask options were relatively insensitive to the challenge aerosol particle size for the range of sizes tested. Therefore, use of the overall effective filtration efficiency for inhalation measured here is reasonable for the Wells-Riley model. It should be noted that the conditions measured here represent an inhalation condition with steady flow, which ignores the unsteadiness of real breathing. However, we anticipate that the differences in filtration efficiency this causes will be moderate. Additionally, the measurements performed here were for inhalation and size-resolved exhalation measurements were not available. We also anticipate that the impact on the filtration that this has should be moderate (although it may be somewhat size dependent).

### 4.3 Mask filtration aerosol dynamics

As a means of testing the assumption that exhalation filtration efficiency performance should behave similar to inhalation, and that the mask fitters perform just as well for exhalation, measurements of the room aerosol concentration versus time were made with the instructor manikin (see Figure 2) emitting aerosol at the front of the room without and with a disposable non-medical procedure and a mask fitter on the instructor manikin. The aerosol flowrate of 20.9 L/min simulated a light to medium exertion breathing rate [61] representative of an instructor lecturing. The aerosol particle size distribution was similar to that shown previously in Figure 4, although the distribution sampled directly from the manikin’s mouth had a smaller count median diameter of 0.222 *µ*m, a smaller geometric mean diameter of 0.286 *µ*m, and a slightly smaller geometric standard deviation of 1.82. The total concentration of aerosol being emitted form the manikin’s mouth was 2.46*×*10^6^ cm*^−^*^3^.

Results for the room aerosol concentration versus time measured just inside the air return for the room using the ELPI with and without the mask and mask fitter on are shown in Figure 16. Well-mixed model estimates were calculated using Equation 7 using the measured particle concentration emitted from the manikin’s mouth along with the measured room and aerosol flowrates and the particle loss rate due to settling determined earlier (*λ_other_* = 0.35 h*^−^*^1^). For the modeled case with the procedure mask and mask fitter, an effective filtration efficiency of 94.9% was used (as measured with the FTM mask fitter).

**Figure 16:**
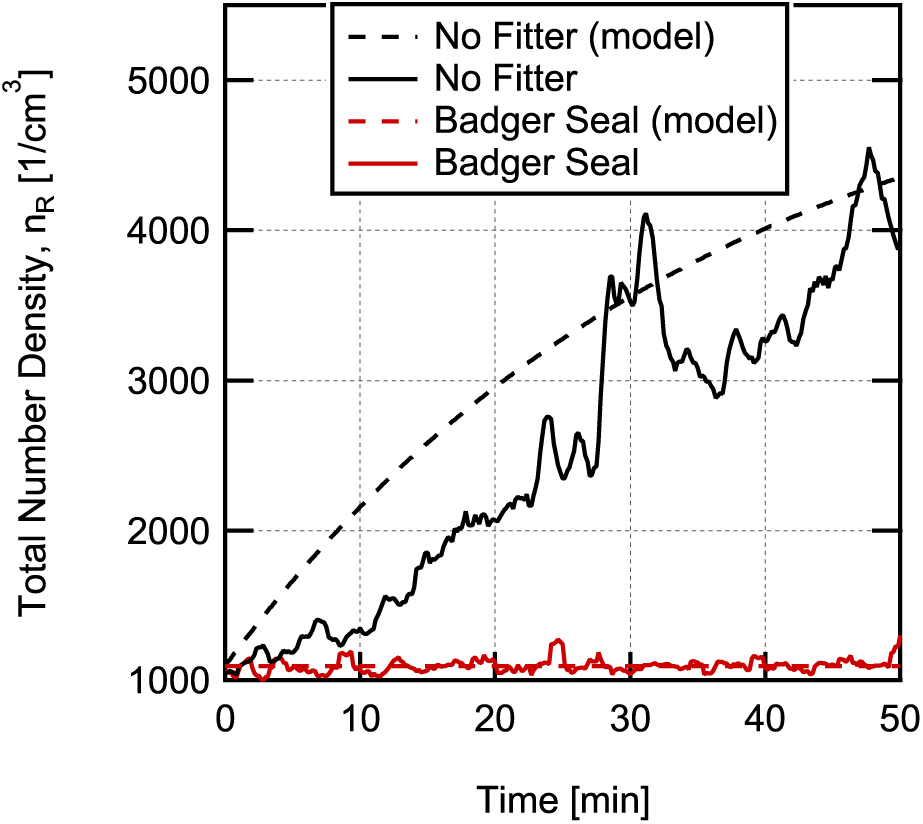
Aerosol number density versus time measured in the room return grille from start of aerosol flow out of manikin’s mouth without and with a single-use procedure mask and Badger Seal mask fitter on the manikin. Results are normalized by the steady-state value that would be achieved without the mask.

It is clear from the results shown in Figure 16, that for exhalation the combination of the single-use procedure mask and the mask fitter significantly reduces the aerosol concentration in the room relative to the case without a mask. The results, when compared with the model calculations, suggest that the mask with fitter provides high effective filtration efficiency for exhalation similar to the 90 to 95% filtration efficiency measured for inhalation, supporting the assertion that exhalation effective filtration efficiencies may be similar to the inhalation effective filtration efficiencies measured (at least for the mask with mask fitter). The results in Figure 16 also illustrate again that the simple well-mixed room control volume model is relatively accurate for capturing the time-evolution of the average concentration of aerosol particles in the room. Deviations between the model and measurements exist, but using measured input parameters, these deviations are generally within a factor of two of the measurements, and the peak concentrations reached appear to be within *±*20% of the measurements. This is much less than the typical uncertainty in quanta emission rate, which can be uncertain by an order of magnitude or more [10, 13].

## 5 Wells-Riley model results and discussion

The purpose of studying the aerosol dynamics and distribution in the classroom and the mask effective filtration efficiencies was to validate the underlying model assumptions and determine accurate input values for the Wells-Riley model. This allows increased confidence in the model’s ability to estimate conditional infection probability.

### 5.1 Model input parameters and assumptions

The aerosol distribution results verified that the well-mixed approximation used in the Wells-Riley model is reasonably accurate (within *∼* a factor of 2) for individuals that are physically distanced from a potentially infected individual by a distance *>* 2-3 m. The aerosol dynamics results provide an estimate of the loss rate of particles due to particle settling and HVAC system losses. Finally, the mask effective filtration efficiency measurements provide effective filtration efficiency with and without a mask fitter for the four different masks tested. Additionally, the time-resolved measurements with a mask on the source manikin confirmed the high effective filtration efficiency obtained with the procedure mask used with a mask fitter for exhalation, in addition to the inhalation measurements.

With the aerosol behavior characterized in the classroom, the results were used as inputs to the Wells-Riley model to assess the conditional infection probability in the event that one of the classroom’s occupants was COVID-19 positive and actively shedding the SARS-CoV-2 virus. The conditional infection probability (*P*), given by the exponential dose response model in Equation 1, represents the probability of infection for any one event (in this case for an in-person class meeting).

We evaluate conditional infection probability for the following three scenarios:

a. Infectious instructor (speaking loudly), susceptible students (sedentary),
b. Infectious student (speaking), susceptible students, and
c. Infectious student (speaking), susceptible instructor (light exercise).

Given the differences in expiratory activity and activity level, different values of quanta emission rate and breathing rate are used for the the instructor and students. Relevant values are provided in Table 5. The breathing rate for the students was chosen to correspond approximately to those from Adams [58] for sitting or standing for adults and approximately match the flow rate used in the mask effective filtration testing. The value used for the instructor corresponds approximately to the breathing rate for walking for adults from Adams [58] (similar values are used in [10]). The quanta emission rate, 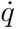, is higher for the instructor who is assumed to be speaking loudly and frequently compared to the students who are, generally, seated and speaking less frequently. Values were determined using the emission rate distributions provided by Buonanno et al. [10] for light activity, speaking loudly, and for light activity, speaking, for the instructor and students, respectively, and then determining the approximate value of emission rate which has an infection probability that is equal to the risk defined by

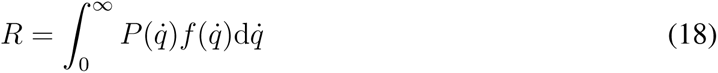

where *R* is the risk of infection, 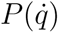 is the quanta emission rate dependent infection probability, and 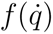 is the probability density function for the quanta emission rate. The risk of infection takes into account the distribution of potential emission rates for an infected individual [10], whereas the infection probability does not. For both emission rates the values chosen in this way correspond to the approximately the 77 percentile of the emission rate distribution, making them representative of a high emission rate scenario.

**Table 5:** Scenarios for infection probability calculations

|  | Scenario A | Scenario B | Scenario C |
| --- | --- | --- | --- |
| Setting | Classroom | Classroom | Classroom |
| Infected Individual | Instructor | Student | Student |
| Emission Rate, $\dot{q}$ | 110 quanta/h | 19.1 quanta/h | 19.1 quanta/h |
| Susceptible Individual(s) | Students | Instructor | Students |
| Susceptible Breathing Rate, $\dot{V}_b$ | 0.540 m<sup>3</sup>/h | 1.38 m<sup>3</sup>/h | 0.540 m<sup>3</sup>/h |

It should be noted that the distributions provided by Buonanno et al. are given in terms of 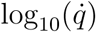 and that to use them appropriately in Equation18 the probability density function should be written as

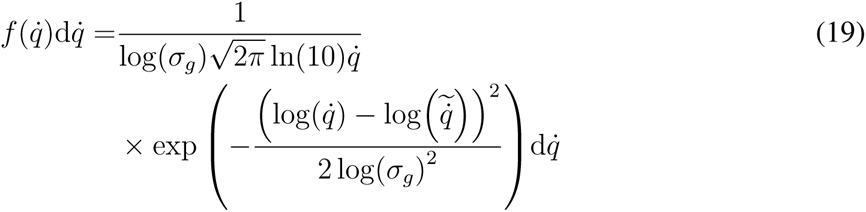

where log is the base 10 logarithm, ln is the natural logarithm, and log(*σ_g_*) and 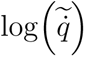 are the distribution parameters called standard deviation and average, respectively, provided in Table 2 of the paper by Buonanno et al. [10].

The room used for probability calculations for all three scenarios was the same classroom used for aerosol dynamics and spatial distribution measurements. The details of the room are provided in Table 6 along with values for the other remaining parameters needed for the calculations. To simplify the estimates, and due to lack of complete information on the filtration capabilities of the air handling unit, it was assumed that all air supplied to the room is fresh air (i.e., quanta free). This will slightly underestimate the infection probability, but due to the relatively high expected filtration efficiency of the air handler filter over the relevant size range (80-90%, similar to a MERV 13 filter [62]) and the large capacity of the air handler resulting in dilution (see Table 1) this should be only a slight underestimation. For simplicity, it is also assumed that time between class periods in the room is long enough such that the initial concentration of quanta in the room is zero. Finally, it is assumed that the mask filtration efficiency for inhalation and exhalation are equal.

**Table 6:**
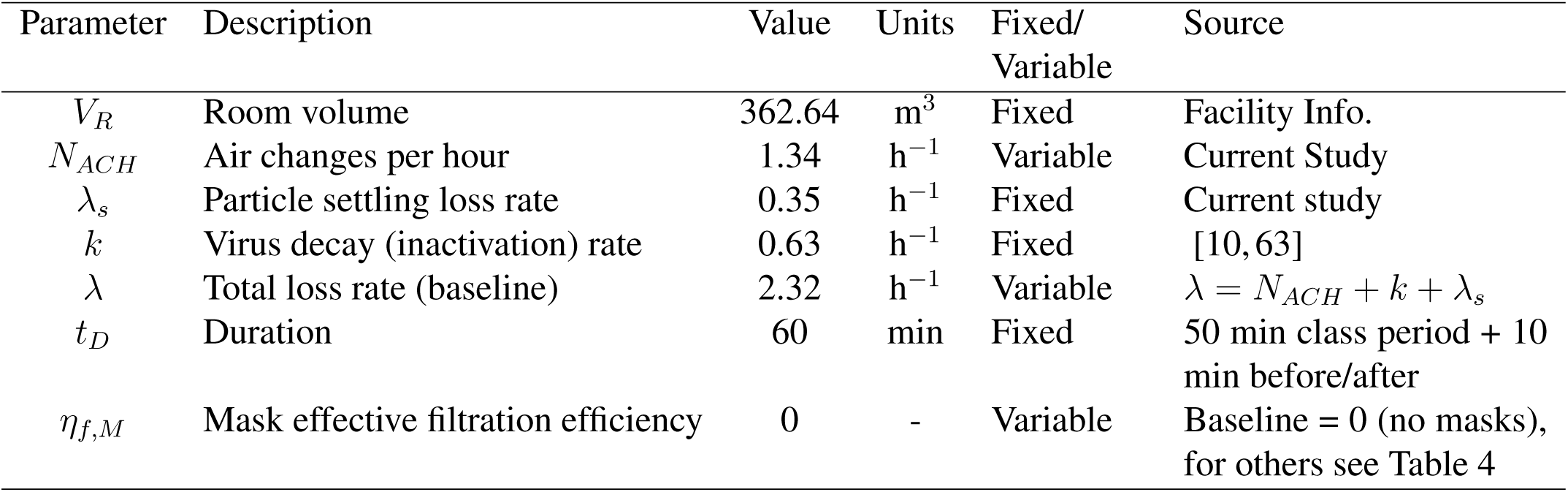
Parameter values for Wells-Riley conditional infection probability estimates. For parameters that are varied in the calculations (variable) values parameter values shown are the baseline values used or determined in the current study.

### 5.2 Model results with varying mask filtration efficiency and ventilation (loss rate)

The conditional probability of infection was predicted (under the condition of one infectious individual being present in the room) for the three scenarios described in Table 5 as a function of mask filtration efficiency with the baseline air exchange rate *N_ACH_* =1.34 h*^−^*^1^ (assuming that everyone in the room is wearing the same type of mask), and as a function of air exchange rate for the case with no one wearing masks, the results for both cases are shown in Figure 17. Of the three different scenarios studied, scenario A, the case with an infected instructor and susceptible students, results in the highest conditional probability due to the assumed higher quanta emission rate for the instructor. Scenario B with an infected student and susceptible instructor had intermediate probability, and scenario C with an infected student and susceptible students has the lowest probability, due to the lower quanta emission rate combined with the lower breathing rate.

**Figure 17:**
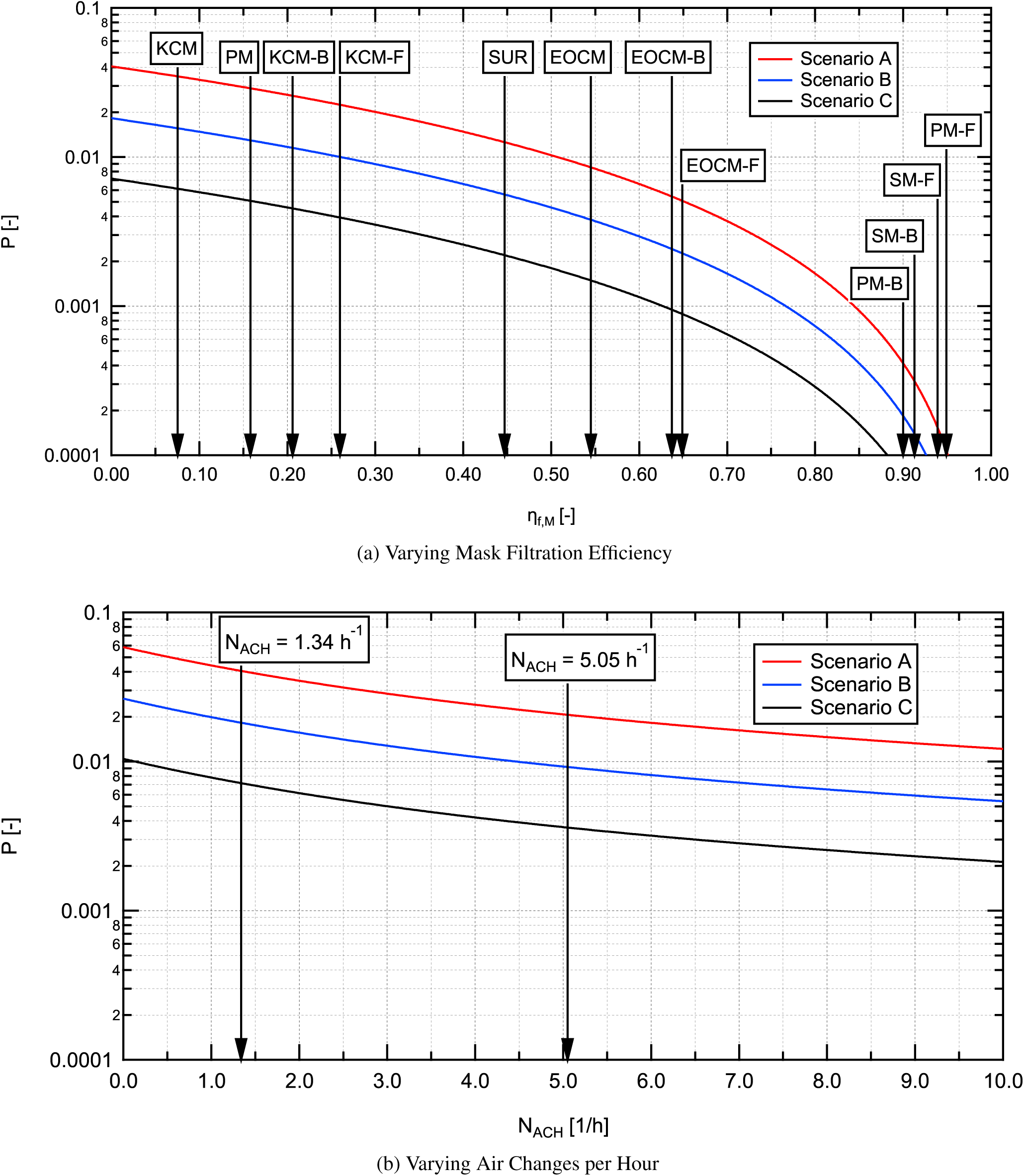
Estimated conditional probability of infection for aerosol (long-range airborne transmission) in a classroom for different intervention scenarios for (a) varying mask filtration efficiency (*η_f,M_*) and (b) varying number of air changes per hour (*N_ACH_*).

The spacing between curves for the three scenarios shown in Figures 17a and 17b remains constant on the vertical log scale as the parameter of interest is varied. This indicates a constant factor between the curves as a result of the differences in quanta emission rate and breathing rates for the three scenarios. As mask filtration efficiency is increased, the conditional infection probability decreases, somewhat slowly at first, then the rate of decrease increases substantially for higher mask effective filtration efficiencies. At a mask effective filtration efficiency of 0.5 (50%) the conditional infection probability is reduced by a factor of 4 relative to the baseline case with no masks (*N_ACH_* =1.34 h*^−^*^1^). With everyone wearing a mask with an effective filtration efficiency of 0.9 (90%), the conditional infection probability is reduced by a factor of 100, as one would anticipate from Equation 15. This is due to the 10x decrease in quanta emitted to the room combine with the 10x decrease in quanta inhaled through a mask, resulting in a total reduction of a factor of 100, greatly reducing the infection probability relative to the baseline value.

For the scenarios as a function of varying air changes per hour (Figure 17b), the curves decrease faster at low values of *N_ACH_* and conditional probability decreases slower with increasing *N_ACH_* at higher values, indicating a diminishing return for increasing ventilation rates. Increasing from the baseline value of *N_ACH_* =1.34 h*^−^*^1^ to 5.05 h*^−^*^1^ results in a decrease in infection probability by about a factor of two. Further increasing the air change rate to 10 h*^−^*^1^ results in an additional factor of 1.7 decrease in infection probability. It is interesting to note that even at an air change rate of 10 h*^−^*^1^ the infection probability for scenario A cannot be reduced below 0.01 or 1%, whereas with everyone wearing the EOC mask without a fitter with the baseline air exchange in the room (*N_ACH_* = 1.34), the probability of infection can be reduced to *<*0.009 (*<*0.9%).

In the results shown in Figure 17, either everyone was assumed to be wearing a mask (Figure 17a) or everyone was assumed to not be wearing a mask (Figure 17b), therefore, it is useful to look at the intermediate case where only some individuals are wearing a mask and compare it to the other cases. The relative change in conditional infection probability as a function of mask effective filtration efficiency, comparing a case with everyone wearing a mask to a case with only the source or susceptible individual wearing a mask, is shown in Figure 18. On this plot, the case of no one wearing a mask would be a straight horizontal line at a value of one. The results illustrate the strong decrease in infection probability achieved if both the infectious and susceptible individuals are wearing masks (everyone wearing a mask). There is a significant decrease in infection probability over the entire range of effective filtration efficiencies from 0.2 to greater than 0.95 for the case with everyone wearing a mask relative to only the susceptible or infectious individuals wearing a mask. Additionally, it is clear that even if others around you are not wearing masks it is beneficial for individuals to wear the best-fitting highest effective filtration efficiency masks available.

**Figure 18:**
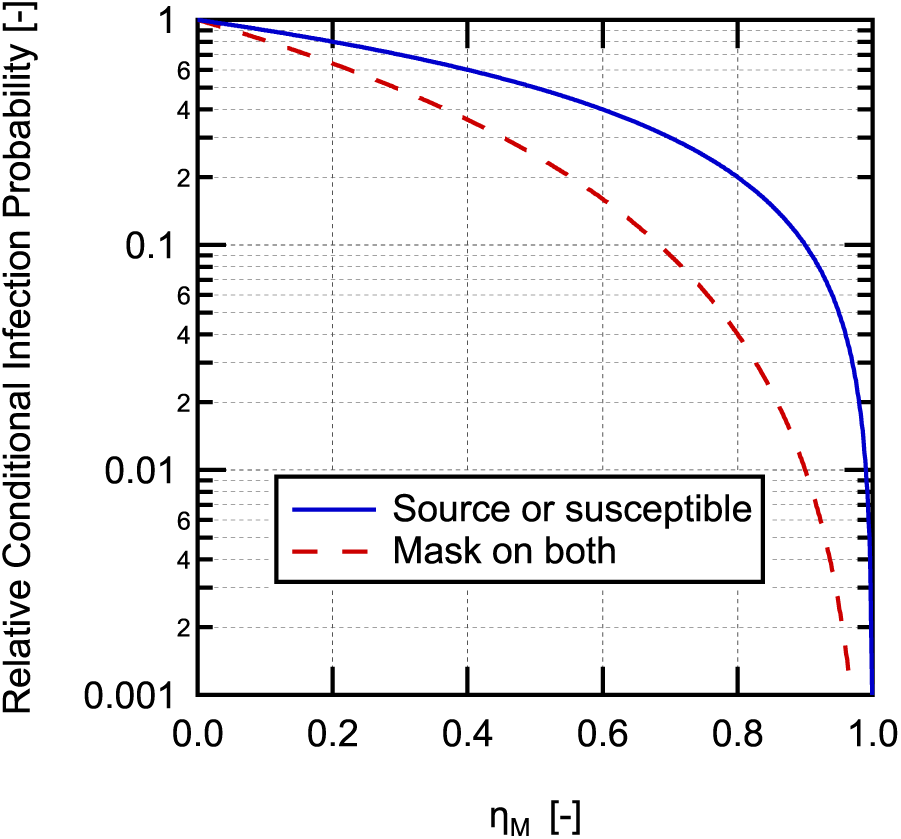
Relative conditional probability of infection (normalized to no mask case) as a function of mask filtration efficiency for a case with only the susceptible individuals or the infectious individual wearing compared to the case with everyone, both the susceptible and infectious individuals wearing the same type of masks.

To better visualize the relative impacts of ventilation and mask effective filtration efficiency, the conditional probability for scenario A was calculated as a function of both of these parameters and plotted as a conditional probability contour map in Figure 19. It should be noted that the left axis in Figure 19 can also be interpreted as total loss rate (*λ*) by increasing the values listed on the axis by *λ_s_* + *k* = 0.98. For the highest values of conditional infection probability, i.e., low mask filtration efficiency and air exchange rate, the contours on the plot are at nearly a 45*^◦^* angle relative to the horizontal axis, indicating that both parameters influence the infection probability in this region. However, as infection probability is reduced (by either increasing *N_ACH_* or increasing *η_f,M_*), the contours become more vertical indicating a reduction in sensitivity to *N_ACH_* when compared to *η_f,M_*. It is also important to recognize again that with ventilation rate alone (for the range studied here) it is not possible to reduce the probability of infection below 0.01, whereas with improved mask effective filtration efficiency, achievable with inexpensive disposal masks combined with a mask fitter, it is possible to decrease infection probability to less than 0.0001 at the baseline ventilation rate, more than two orders of magnitude decrease in infection probability.

**Figure 19:**
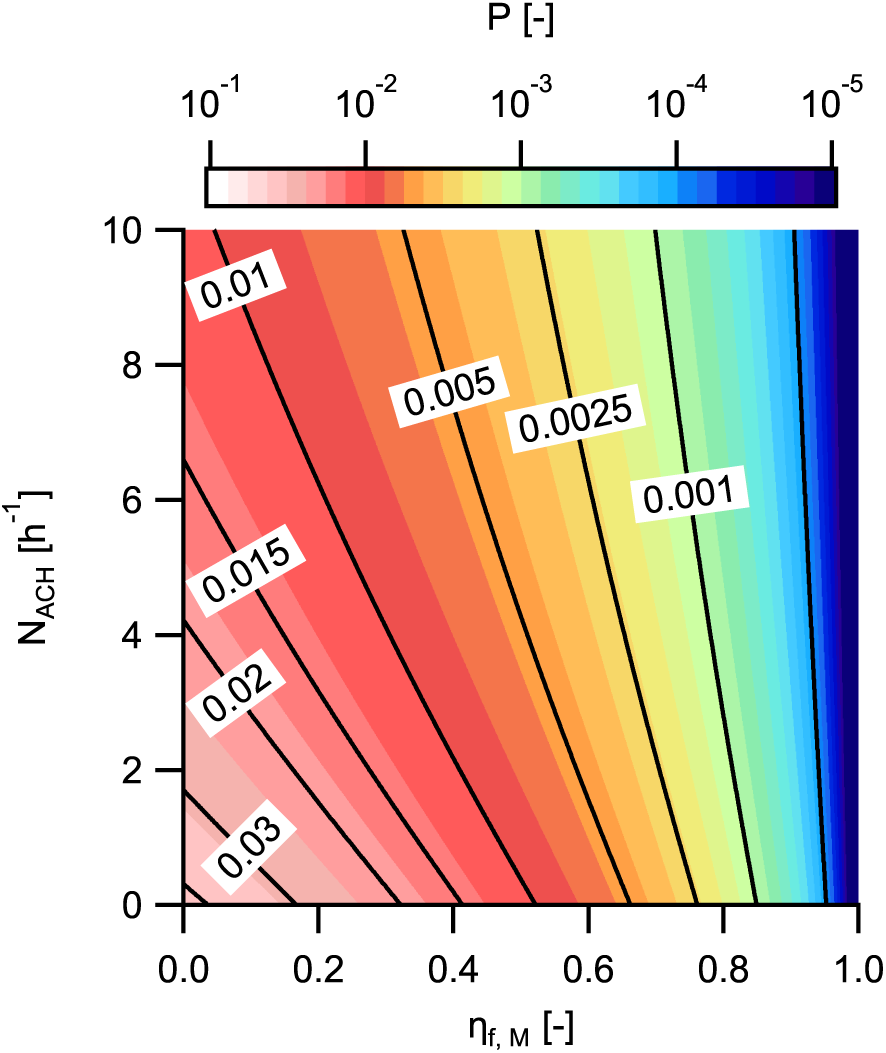
Conditional probability map with contours overlaid as a function of air changes per hour and mask effective filtration efficiency for Scenario A.

### 5.3 Results for specific interventions

Conditional infection probabilities for the three scenarios in Table 5 were also estimated for specific combinations of HVAC flowrate (air changes per hour) and masks worn by students and an instructor. Three values of HVAC flowrate were used: 1.34 ACH (Baseline), 5 ACH, and 10 ACH. The use of an in-room recirculating air purifier could be considered as part of the HVAC flowrate in these calculations assuming 100% filtration efficiency for the purifier and no impact on the well-mixed model approximation. Calculations were performed for cases with everyone in the room wearing knit cotton cloth masks (KCM), the EOC mask (EOCM), and the inexpensive procedure mask (PM) both with and without use of mask fitters (filtration values used are those for the masks with the FTM fitter). The surgical mask was not considered here due to higher cost, low availability, and the desire not to impact supplies for medical professionals, but performance would be similar to the EOCM without a fitter and almost identical to the procedure mask with a mask fitter. An additional case is also considered where the instructor is wearing a procedure mask with a mask fitter and the students are all wearing cloth masks. This case is focused on source control of the person who will likely have the highest quanta emission rate due to their known activity and who could be trained to consistently and properly don a mask and mask fitter. For each combination of interventions, the three scenarios described in Table 5 are calculated corresponding to: (A) an infectious instructor and susceptible students, (B) an infectious student and a susceptible instructor, and (C) an infectious student and susceptible students.

The conditional probability results for the different scenarios and intervention combinations are shown in Figure 20. We have included a dashed line at a conditional probability of 0.001 on the plot for reference, this value was suggested in the recent publication of Buonanno et al. [10] as a target. For the current case, use of this value for the conditional infection probability target gives a likely number of cases (if an infectious individual were present) of *N* = *PN_s_* = 0.001 *×* 16 = 0.016 or an attack rate of 1.6%.

**Figure 20:**
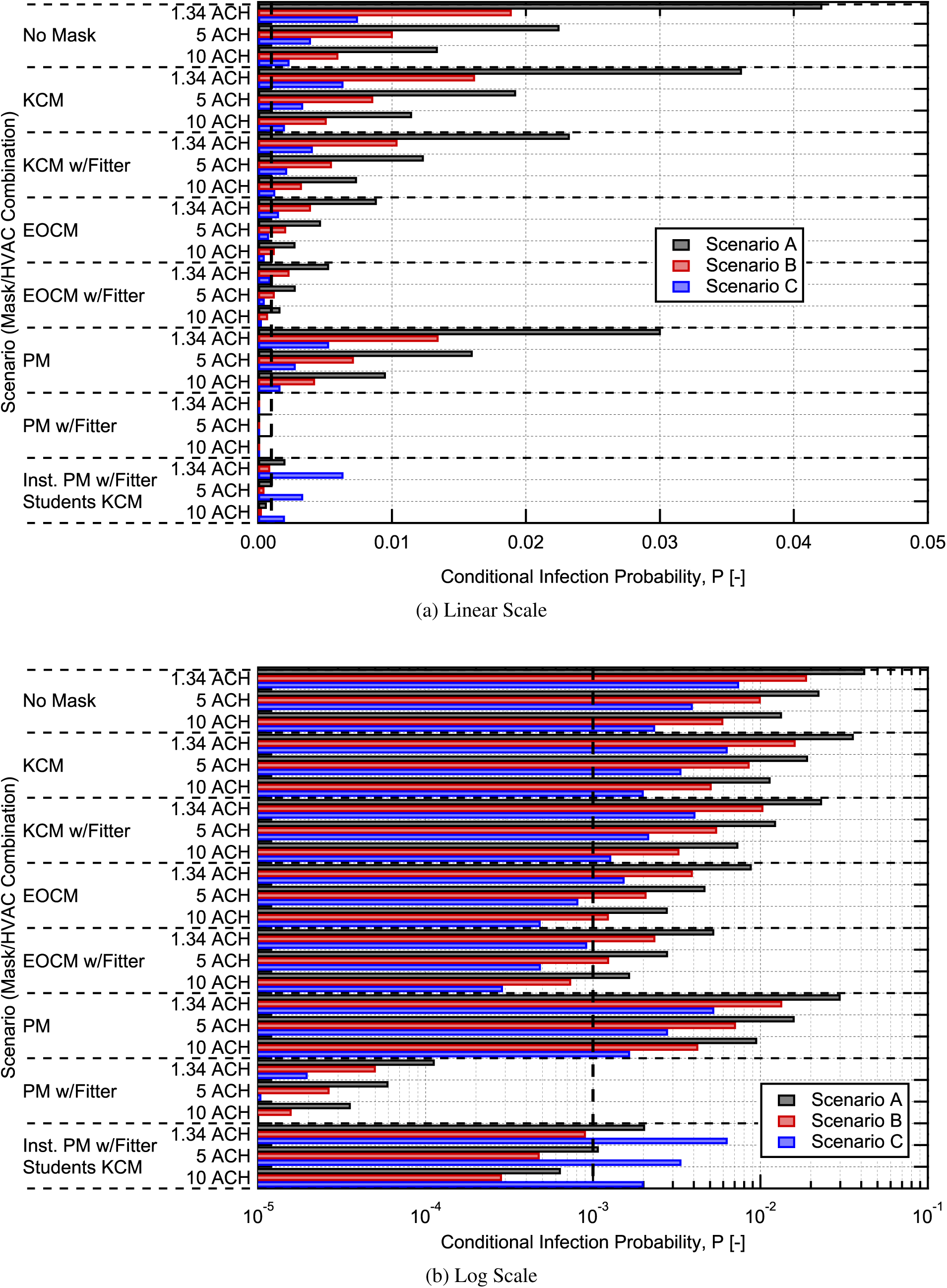
Estimated conditional probability of infection for aerosol (airborne transmission) in a classroom for different interventions combinations, results are shown versus both (a) a linear scale and (b) a log scale. A vertical dashed line is shown at the 0.001 conditional probability level. The mask abbreviations used on the graphs are the same as those defined in Table 4.

For the results in Figure 20, the addition of knit cotton cloth masks for all classroom occupants yielded a modest 15% reduction in the conditional probability of aerosol infection compared to the baseline with no masks. In comparison, the protective measure of increasing the ventilation rate from 1.34 ACH to 5.0 ACH resulted in almost a factor of two (1.87x) reduction in conditional probability, regardless of mask worn. If a “fitter” is added to help seal the knit cotton mask to the user’s face, the conditional probability of infection is reduced by almost a factor of 2 (1.81x) relative to the no mask baseline. This reduction is similar to increasing the ventilation rate from 1.34 to 5.0 ACH. This indicates that well fit cloth masks can play a significant role in reducing infection probability.

Combining higher ventilation rates with masks is synergistic and the impacts are multiplicative as indicated in Equation 15. For example, the combination of knit cotton masks with mask fitters and an increase in ventilation rate from 1.34 to 5.0 ACH results in a factor of 3.4 reduction in conditional infection probability relative to the no mask baseline, which is equal to the product of the reductions for each individually. It is interesting to note that this combined reduction is greater than could be achieved by increasing HVAC flow from 1.34 to 10 ACH.

As we move to masks that have higher effective filtration efficiencies, we see more dramatic reductions in the conditional infection probability. For a moderate efficiency reusable mask like the EOC mask, factors of 4.75 and 8.2 reduction can be achieved without and with the use of a mask fitter, respectively. For the high efficiency disposable procedure mask, the fit was poor without a mask fitter, and it had only a 15.8% effective filtration efficiency, but with a mask fitter an effective filtration efficiency of 94.8% could be achieved resulting in almost a factor of 400 reduction (371x) in conditional infection probability. This demonstrates the significant potential for reduction in aerosol transmission of SAR-CoV-2 achievable with relatively inexpensive masks and mask fitters.

In addition to the EOC mask whose results were shown here, a different homemade 3-ply spunbond polypropylene (Agfabric 1.5 oz floating row cover) mask was also tested. The mask pattern was similar to the pattern for the EOC mask. This mask had a measured effective filtration efficiency of 65.2*±*2.7% without use of a mask fitter providing a reduction in infection probability of 8.2x, similar to the EOC mask with the mask fitter, when worn by all students and the instructor relative to the no mask baseline. The level of filtration this mask and the EOC mask achieved without a mask fitter (on the order of 55-70% filtration efficiency) is a good target for common face coverings as it provides significant infection probability reduction for aerosol transmission with a reusable mask, made of a material that is widely available, has low pressure drop, and good filtration characteristics, i.e., a high filter quality [19, 53].

The case with the instructor using a procedure mask and mask fitter with the students using knit cotton masks, provides a look at how source control can provide significant reductions in infection probability and is attractive due to its cost effectiveness. In this case, by using a high efficiency mask on the person who is likely the highest emitter due to their known activity level, a reduction in the maximum conditional infection probability by a factor of 5.67 is achieved. Therefore, targeted interventions, where one or a few individuals are performing activities known to have high quanta emission rates, can have a large impact on infection probability. It also faces the reality that equipping and training an instructor on the proper donning of their mask is more readily achievable than relying on the same administrative control strategy for a larger number of student occupants. The conditional infection probability results for the different interventions illustrate a few important points. First, increasing HVAC flowrate (ventilation), when possible, and ensuring a minimum acceptable ventilation rate is important for reducing infection probability, but there is a decreasing return on investment, as seen previously in Figure 17. Second, the three scenarios (from Table 5) result in significantly different probabilities due to the differences in emission rate and breathing rate between them. For the infectious instructor, susceptible students case (Scenario A), the conditional probability of infection is the highest due to the highest emission rate assumed for the lecturing instructor. For Scenario C the quanta emission rate is a factor of 5.76 lower (breathing rate is the same) and the conditional probability drops by a factor of 5.65, i.e., approximately proportional with the reduction in quanta emission rate as expected based on Equation 15 (the results shown here can easily be scaled to make estimates for different number of infectious individuals in the room, quanta emission rates, loss rates, room volumes, breathing rates, and mask filtration efficiencies). This suggests that source control for cases where high quanta emission rates are likely is a potentially effective strategy. Finally, it is clear that masks and the use of mask fitters to reduce mask leakage (or better mask design to achieve the same result) can provide significant reductions in infection probability for aerosol transmission greater than those easily achievable by ventilation alone. For example, the use of the procedure mask with a mask fitter by everyone in the room enables conditional infection probabilities of 0.0001 or lower. However, the high material filtration efficiency procedure mask used without a mask fitter provides only a moderate reduction (*∼*30%) in infection probability due to its poor fit.

From the results shown, it is clear that ventilation rate alone will not be sufficient to achieve conditional infection probabilities below 0.001 in the classroom space considered for this study. Thankfully, as Equation 15 and the results shown illustrate, the combined effects of masks and ventilation are synergistic (multiplicative) and use of both simultaneously results in greater reductions in infection probability. For example, use of moderate filtration efficiency masks like the EOC mask along with elevated HVAC flowrates can come close to achieving 0.001 conditional infection probability in 2 out of the three scenarios (B and C).

## 6 Summary and Conclusions

Modeling of infection risk and infection probability using a simplistic well-mixed model with an exponential dose assumption provides useful guidance on the efficacy of interventions to reduce aerosol transmission infection probability in classroom settings during the ongoing COVID-19 pandemic. In this work, we directly evaluated a number of the required input parameters for the 0-D concentration evolution model and assess the validity of the well-mixed model assumption in a classroom setting with mechanical ventilation. This information was then used in the Wells-Riley model conditional infection probability estimates for SARS-CoV-2 in the same classroom setting to compare the effectiveness of interventions and combinations of interventions to reduce conditional infection probability: ventilation rate, and mask effective filtration efficiency.

Testing of aerosol dynamics and distribution along with mask effective filtration efficiency was performed by seeding a classroom at the University of Wisconsin-Madison with NaCl aerosol and sampling aerosol concentrations in the room and through masks installed on a manikin. The time-resolved measurements of aerosol dynamics showed an exponential rise when seeding was turned on and an exponential fall when seeding was turned off. Concentrations measured in the front and back of the room when seeding from a single central location showed aerosol concentrations within a factor of two during the increase in aerosol concentration and were almost exactly equal after seeding was turned off. The rise and fall of concentration in the room were fit to the expected exponential time dependence and a total loss rate coefficient (inverse time constant) was extracted. The extracted loss rates were slightly larger than the air change rate in the room, indicating the presence of other loss mechanisms such as particle settling in the room and in the duct work, and providing a quantitative estimate of the average loss rate. Aerosol distribution measurements were performed after the room concentration had reached steady state. The aerosol size-resolved spatial distribution was dependent on whether the simulated infected student manikin was wearing a mask or not wearing a mask. When not wearing a mask, more penetration of aerosol in the direction the manikin was facing was observed and concentrations up to 60% higher than the minimum in the room were observed at a distance 3 m in front of the manikin. For the case with a mask present elevated aerosol concentrations were limited more to the region within 2.5 m of the manikin. Some size dependence was also observed in the non-uniformity, likely resulting from a size-dependent settling loss rate.

The effective filtration efficiency for inhalation of masks installed on a manikin was measured with and without the use of two different mask fitters designed to help seal the mask to the user’s face. Two of the masks tested were washable reusable masks, one made of 4-layers of knit cotton and the other of 3-layers of non-woven spunbond polypropylene. The other two masks were disposable, one was a common single-use disposable mask (termed procedure mask throughout) and the other was an ASTM F2100 level-2 surgical mask. The effective filtration efficiency provides information on the efficacy of masks as installed on the user’s face. The results demonstrated the importance of mask fit with poorly fitting masks showing significantly degraded filtration efficiency (in the worst case decreasing filtration efficiency by a factor of *∼*6). The results also demonstrated that the knit-cotton mask, even when using a fitter, was only able to achieve an effective filtration efficiency of 26%, whereas the polypropylene mask (EOC mask) was able to achieve effective filtration efficiencies of up to 65%. Use of a mask fitter was shown to greatly improve the effective filtration efficiency of the masks. The exception to this was the spunbond polypropylene mask (EOC mask) which was able to achieve a good fit without the use of a mask fitter. Use of the surgical mask or procedure mask with a mask fitter enabled effective filtration efficiencies approaching 95%.

Conditional probability estimates were performed for the same classroom as the measurements. The results illustrated that both ventilation rate and mask effective filtration efficiency decrease the long-range airborne probability of infection for SARS-CoV-2. However, the use of air exchange rate alone was not able to provide greater than approximately a factor of 4 reduction in infection probability, for ventilation rates up to 10 h*^−^*^1^. On the other hand, mask effective filtration efficiency can be used to achieve reductions in infection probability greater than a factor of 100. Even for masks with a modest effective filtration efficiency of *∼*55%, achievable with reusable spunbound polypropylene mask without a mask fitter, *>* 4x reductions in infection probability can be achieved. These results emphasize the importance of ventilation and masks for reducing aerosol transmission and the synergistic nature of both interventions.

In this work, masks were installed on the manikins in a manner that achieved the best expected fit for the mask when not using a mask fitter. In reality, masks worn by users in public are often worn in a manner where even more leakage around the mask occurs, due either to poor mask design or due to poor user adjustment, resulting in low effective filtration efficiency. To achieve the reductions in infection probability estimated here for mask wearing, significant education of the public is required on proper mask wearing and on the substantial reductions in infection probability for the mask user. Just recently, the CDC has acknowledged that wearing a mask also protects the wearer [37], but the message is still not clearly communicated to the majority of the public. This is an area of public education that may significantly benefit local municipalities.

The major conclusions from this work are summarized here:

1. Particle loss rates for the classroom determined by measurements of air exchange rate for the room match well with those determined from aerosol concentration time evolution in the class room, giving improved confidence in inputs to the well-mixed model.
2. Based on *in situ* measurements in a representative classroom with forced air mechanical ventilation, the well-mixed assumption holds to a reasonable approximation (within better than a factor of 2) for the classroom space studied.
3. Leakage rates around masks are typically in the range of 20-80% when fit reasonably well to the user’s face, resulting in effective filtration efficiencies that are much lower than the material filtration efficiency.
4. With the use of a mask fitter, one can achieve an effective filtration efficiency approaching the material filtration efficiency for a mask. Inexpensive disposable masks used with a mask fitter can achieve approximately 95% effective filtration efficiency in some cases.
5. Ventilation of spaces with fresh air or well-filtered air is important, ventilation rates greater than *>* 5 ACH result in only modest additional reductions in infection probability and generally are not merited due to their high cost, energy usage, and increase in noise and room air velocities.
6. With all occupants in the indoor classroom masks, long-range airborne conditional infection probabilities are greatly reduced and reductions of 4x, 10x, or even 100x can be achieved.
7. In certain scenarios, source control, i.e., providing an individual performing a known high emission rate activity with a high effective filtration efficiency mask (using a mask fitter), may be an effective approach to limit long-range airborne infection probabilities.
8. Use of masks and ventilation simultaneously are synergistic (providing multiplicative reductions) and together can provide greatly reduced aerosol transmission infection probability.

The conclusions provided here give guidance on the relative effectiveness of ventilation and mask wearing for decreasing long-range airborne infection probabilities. The results specifically emphasize the large potential for effective mask wearing and improved masks, or use of mask fitters, to greatly reduce infection probabilities.

## Supporting information

Supporting Information Fit Testing

## Data Availability

Data that support the findings of this study are available from the corresponding author upon reasonable request.

## 7 Acknowledgements

This work was funded in part by the Emergency Operations Committee at the University of Wisconsin-Madison.

The authors would like to acknowledge the useful inputs and assistance from James Morrsion, Jesse Decker, Jessica Cebula, and Christopher Strang, and others in Environment, Health & Safety, the College of Engineering, and Facilities Planning and Management at the University of Wisconsin-Madison.

The authors would like to also acknowledge Lennon Rodgers and employees of the University of Wisconsin-Madison Makerspace for their efforts on the design and manufacturing of the Badger Seal mask fitter which was inspired in part by the initial portions of this work.

The authors would also like to thank graduates students Logan Kossel and James Rice as well as Prof. Alejandro Roldán-Alzate for their useful inputs and assistance.

## 8 Conflict of interest

The authors of this paper certify that they have NO affiliations with or involvement in any organization or entity with any financial or non-finanical interest in the subject matter or

## 9 Author contributions

**David Rothamer:** Conceptualization (equal); Data curation (lead); Formal Analysis (lead); Investigation (lead); Methodology (lead); Project administration (lead); Resources (equal); Software (lead); Supervision (lead); Validation (lead); Visualization (lead); Writing - Original draft preparation (lead); Writing - review and editing (equal); **Scott Sanders** Conceptualization (equal); Investigation (supporting); Methodology (supporting); Project administration (supporting); Resources (equal); Supervision (supporting); Writing - review and editing (equal); **Tim Bertram:** Conceptualization (supporting); Investigation (supporting); Methodology (supporting); Writing - review and editing (equal); **Douglas Reindl:** Conceptualization (supporting); Investigation (supporting); Methodology (supporting); Writing - Original draft preparation (supporting); Writing - review and editing (equal);

## Notes

### Competing Interest Statement

The authors have declared no competing interest.

### Author Declarations

No approval IRB approval required for this research

