## Supporting Information Fit Testing for "Strategies to minimize SARS-CoV-2 transmission in classroom settings: Combined impacts of ventilation and mask effective filtration efficiency"

December 31, 2020

### 1 Fit Testing

#### 1.1 Fit Test Setup

We also performed fit testing on various masks, with and without mask fitters, using a commercial fit tester (TSI PortaCount Pro+ 8038). Fit test measurements were performed by a single researcher (one of the authors) that donned the masks and recorded the data in a room with closed doors. A commercial particle generator (TSI 8026) was used to increase the particle number density to approximately 4 times the ambient level, based on fit tester readings. Particle size distributions for such scenarios typically peak near 40 nm [1]. For each mask tested, complete fit tests were performed, and the fit tester was also operated in Real Time mode for 120 seconds while the researcher was reciting aloud the standard fit test language. The results from both testing modes were consistent, but fit test results reported here are exclusively from Real Time mode tests. The results are reported as effective filtration efficiency values, where the error bars represent the range of effective filtration efficiencies observed during a test.

#### 1.2 Fit Test Results

The effective filtration efficiencies recorded during fit testing are shown in Figure 1. The error bars represent the variability observed during the 120-s duration test which was performed in Real Time mode. For the well-adjusted fitter cases, the researcher optimized the fitter adjustment using only a mirror (instrument data was not used to adjust the fitter). For the poorly-adjusted fitter cases, the fitter was purposely adjusted to allow a noticeable leak on the left side of the researcher's face and another leak on the right side of the wearer's nose. A picture of the Original Badger Seal shown in Figure 2a and the Ear Loop Badger Seal is pictured in Figure 2b; both are detailed at https://making.engr.wisc.edu/mask-fitter/.

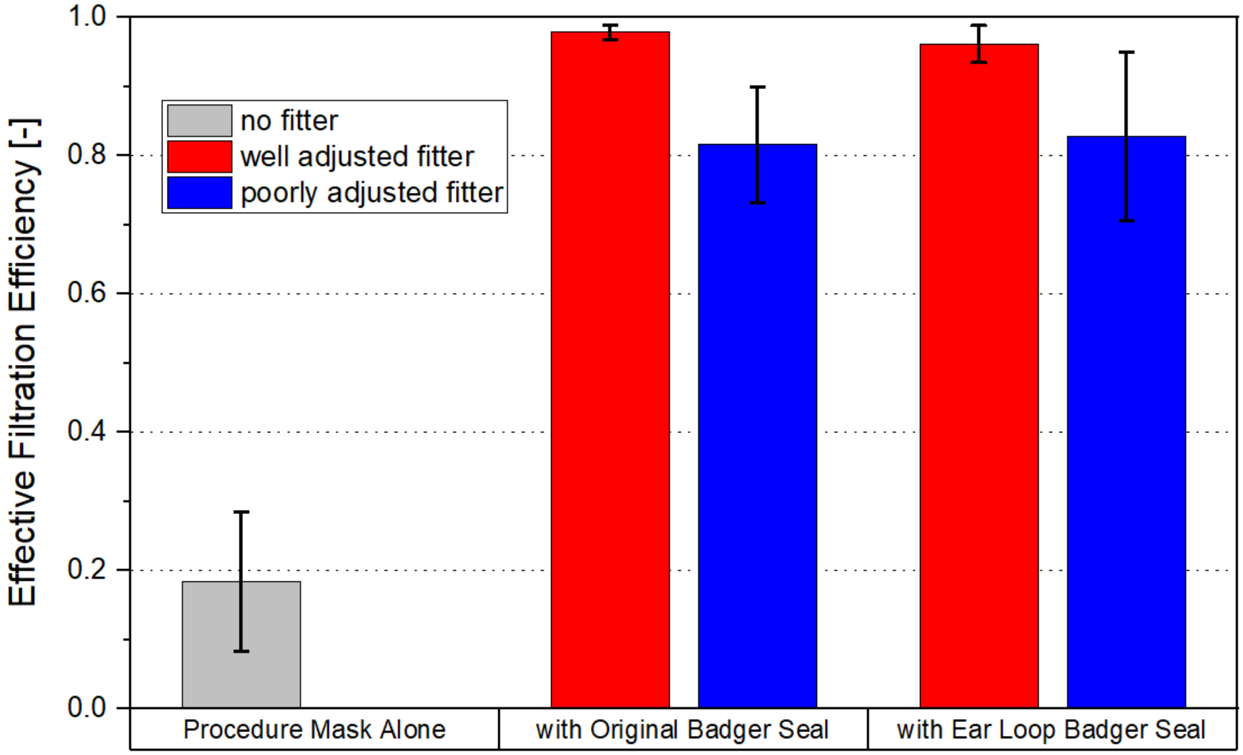


Figure 1: Fit test results with the procedure mask alone and with the Badger Seal Mask fitter both well adjusted and poorly adjusted.

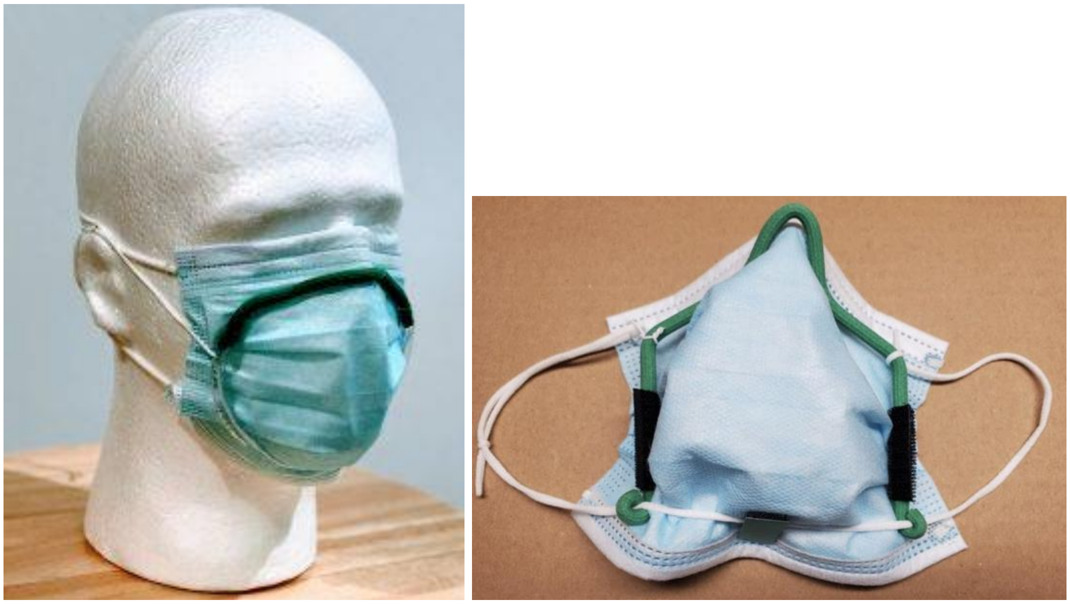

(a) Original Badger Seal (b) Ear Loop Badger Seal

Figure 2: (a) Image of the Original Badger Seal and (b) Image of the Ear Loop Badger Seal version.
